# Homologous and heterologous boosting with CoronaVac and BNT162b2: a randomized trial (the Cobovax study)

**DOI:** 10.1101/2022.08.25.22279158

**Authors:** Nancy H. L. Leung, Samuel M. S. Cheng, Carolyn A. Cohen, Mario Martín-Sánchez, Niki Y. M. Au, Leo L. H. Luk, Leo C. H. Tsang, Kelvin K. H. Kwan, Sara Chaothai, Lison W. C. Fung, Alan W. L. Cheung, Karl C. K. Chan, John K. C. Li, Yvonne Y. Ng, Prathanporn Kaewpreedee, Janice Z. Jia, Dennis K. M. Ip, Leo L. M. Poon, Gabriel M. Leung, J. S. Malik Peiris, Sophie A. Valkenburg, Benjamin J. Cowling

**Affiliations:** WHO Collaborating Centre for Infectious Disease Epidemiology and Control, School of Public Health, LKS Faculty of Medicine, The University of Hong Kong, Pokfulam, Hong Kong Special Administrative Region, China; Laboratory of Data Discovery for Health; Hong Kong Science and Technology Park, New Territories, Hong Kong Special Administrative Region, China; HKU-Pasteur Research Pole, School of Public Health, Li Ka Shing Faculty of Medicine, The University of Hong Kong, Pokfulam, Hong Kong Special Administrative Region, China; Centre for Immunology and Infection; Hong Kong Science and Technology Park, New Territories, Hong Kong Special Administrative Region, China; Department of Microbiology and Immunology, Peter Doherty Institute of Infection and Immunity, The University of Melbourne, Melbourne, Victoria, Australia

## Abstract

**Background:** There are few trials comparing homologous and heterologous third doses of COVID-19 vaccination with inactivated vaccines and mRNA vaccines.

**Methods:** We conducted an open-label randomized trial in adults >=18 years of age who received two doses of inactivated vaccine (CoronaVac) or mRNA vaccine (BNT162b2) >=6 months earlier, randomised in 1:1 ratio to receive a third dose of either vaccine. We compared the reactogenicity, immunogenicity and cell-mediated immune responses, and assessed vaccine efficacy against infections during follow-up.

**Results:** We enrolled 219 adults who previously received two doses of CoronaVac and randomised to CoronaVac (“CC-C”, n=101) or BNT162b2 (“CC-B”, n=118) third dose; and 232 adults who previously received BNT162b2 and randomised to CoronaVac (“BB-C”, n=118) or BNT162b2 (“BB-B”, n=114). There were more frequent reports of mild reactions in recipients of third-dose BNT162b2, which generally subsided within 7 days. Antibody responses against the ancestral virus, Omicron BA.1 and BA.2 subvariant by surrogate neutralization and PRNT_50_ were stronger for the recipients of a third dose of BNT162b2 over CoronaVac irrespective of prior vaccine type. CD4^+^ T cells boost only occurred in CoronaVac-primed arms. We did not identify differences in CD4^+^ and CD8^+^ T cell responses between arms. When Omicron BA.2 was circulating, we identified 58 infections with cumulative incidence of 15.3% and 15.4% in the CC-C and CC-B (p=0.93), and 16.7% and 14.0% in the BB-C and BB-B arms, respectively (p=0.56).

**Conclusions:** Similar levels of incidence of infection in each arm suggest all third dose combinations may provide similar degrees of protection against prevalent Omicron BA.2 infection, despite very weak antibody responses to BA.2 in the recipients of a CoronaVac third dose. Further research is warranted to identify appropriate correlates of protection for inactivated COVID-19 vaccines.

## INTRODUCTION

The accrual of population immunity to COVID-19 may ultimately allow communities to return to normal, or at least to a “new-normal”. Immunity can be acquired through surviving infections or, preferably, by vaccination. An unprecedented global effort has led to the rapid development and deployment of COVID-19 vaccines.^1^ Vaccines against COVID-19 have generally been developed initially as either a single dose, or two doses of the same vaccine technology platform i.e. a homologous two-dose regimen.^1^ The emergence of variants of concern (VOCs)^2^ and observed decrease in vaccine-induced immune responses within a few months after vaccination^3,4^ have led to the implementations of third dose (“booster”) vaccination programs.^5^ While some booster campaigns have encouraged individuals to receive a homologous third dose, a third/booster dose with a different vaccine platform, i.e., heterologous vaccination, could be more feasible in some locations.^6-8^ There is also a possibility that combining vaccine doses using different vaccine platforms by heterologous prime-boost strategies might enhance the immune response.^9^ Heterologous vaccination has been investigated by a number of clinical trials with mixed results depending on the initial platform and sequence of vaccination.^7,10,11^ Conversely, considerations over adverse events may prompt individuals to prefer one vaccine platform over the other.^8^

In Hong Kong, a subtropical city in southern China, the COVID-19 vaccination programme started in February 2021. The programme initially offered each adult in Hong Kong a choice of two doses of the inactivated whole-virion vaccine CoronaVac (Sinovac) 28 days apart, or two doses of mRNA vaccine BNT162b2 (BioNTech/Fosun Pharma, marketed by Pfizer outside China) 21 days apart, with broad eligibility criteria and availability. Subsequently, healthy adults have been recommended to receive a third dose of either vaccine (i.e., allowing cross-over) as soon as 90 days after the second dose.^8^

Safety, immunogenicity and efficacy are the main aspects evaluated before vaccines are licensed. There are few trials^12^ comparing the potential advantages of homologous or heterologous third doses in individuals who have previously received two doses of an inactivated vaccine or two doses of an mRNA vaccine. Here, we conducted a randomized trial to compare the immunogenicity and reactogenicity to BNT162b2 or CoronaVac in individuals that had previously received two doses of those vaccines. In addition, in early 2022 during the biggest local (fifth) wave the antigenically distinct Omicron BA.2 variant was the major circulating strain,^13^ which provided us an opportunity to evaluate the comparative efficacy of these different vaccine combinations against infection.

## METHODS

### Trial design and participants

This study is an open-label randomized trial conducted in Hong Kong to measure the vaccine (humoral) immunogenicity and reactogenicity of third doses with an mRNA vaccine (BNT162b2) or an inactivated vaccine (CoronaVac) in adults who have previously received two doses of either vaccine. BNT162b2 is a nucleoside-modified mRNA encoding the trimerized SARS-CoV-2 spike glycoprotein formulated in lipid nanoparticles, and each 0.3mL dose contains 30µg of mRNA. CoronaVac is a Vero cell-based, aluminium hydroxide-adjuvanted, β-propiolactone-inactivated vaccine, and each 0.5 ml dose includes 600SU of inactivated SARS-CoV-2 virus.

Individuals were eligible for this study if they were Hong Kong residents, ≥18 years of age at enrolment and had previously received a homologous two-dose primary series of either BNT162b2 or CoronaVac, with the second dose received at least 6 months (180 days) earlier. Exclusion criteria included: a history of confirmed SARS-CoV-2 infection, a delay of ≥43 days between the first two vaccine doses, contraindication for COVID-19 vaccination such as severe allergies, use of medication that could impair the immune system in the last 6 months (except topical steroids or short-term oral steroids), use of immunoglobulins and/or any blood products within 90 days prior to enrolment, and pregnant or nursing, or planning to become pregnant. The study protocol (Clinicaltrials.gov NCT05057169) was approved by the Institutional Review Board of the University of Hong Kong (ref: UW 21-492).

### Randomization and masking

The random allocation process was concealed from both participants and the field investigation team, by way of pre-assignment based on a computer-generated sequence of random numbers administered by clicking a randomization button in the online data collection tool REDCap.

Participants received two-dose CoronaVac or BNT162b2 were randomised separately to receive a third dose of either vaccine in a 1:1 ratio. We used a block randomization structure with block sizes of 2, 4 and 6. The sequence of random numbers was generated prior to the start of the study by a statistician using the computer software package R. For logistical reasons individuals were randomized at the time vaccination appointments were made, and there were dropouts after randomization and before vaccination. However, both the participants and field investigation team were not aware of the intervention until after vaccination, as the vaccination was done by separate unblinded nurses at the community vaccination centres. After vaccination, participants were provided with vaccination cards as required by the local government indicating which vaccine they had received, i.e. participants and field investigation team were also unblinded from this time onwards. The staff conducting laboratory tests were blinded to the third dose allocation.

### Procedures

Participants were enrolled from the general community (see **Supplementary Information** for additional information on enrolment and follow-up). We collected 20ml blood samples immediately prior to vaccination (Day 0) and scheduled follow-up blood draws at 28, 182 days and 365 days after vaccination. In a voluntary subset of approximately 35% of participants we collected additional 10ml whole blood samples at day 0, 7 and 28 for analysis of cell-mediated immune responses (CMI). At each encounter for a blood draw we offered a HKD100 (USD13) incentive in the form of a supermarket gift voucher. After vaccination, participants were provided with vaccination cards and tympanic thermometers, observed for 15-30 minutes for immediate events, and then provided with an e-diary and requested to record daily on possible (delayed) adverse events or symptoms for 7 days as well as any medical encounters. From March 2022, all participants were invited to participate in active surveillance to identify COVID-19 and influenza virus infection using rapid antigen tests (RAT) at home every four days. We also ascertained whether any participants had an infection identified by RAT or PCR after enrolment but prior to the start of active surveillance. The active surveillance is continuing with information collected up to May 31 included in the present analyses.

### Outcomes

The primary outcome measure is the immunogenicity at 28 days after the third dose of either BNT162b2 or CoronaVac, measured as geometric mean titer (GMT) of SARS-CoV-2 serum neutralising antibodies using plaque reduction neutralization test (PRNT_50_). The secondary outcome measures included i) the geometric mean fold rise (MFR) of SARS-CoV-2 serum neutralising antibody titers from baseline (Day 0) to Day 28, ii) magnitude and phenotype of vaccine-specific IFNγ^+^CD4^+^ and IFNγ^+^CD8^+^ T-cell responses at Day 7 and 28, iii) the incidence of solicited local and systemic adverse events, and iv) COVID-19 infections after receiving the third vaccine dose.

### Sample size justification

For the primary outcome of vaccine immunogenicity, i.e. the GMT of SARS-CoV-2 serum neutralising antibodies against the vaccine strain (ancestral virus) at 28 days after vaccination, a sample size of 100 individuals in each study arm was chosen. Based on our preliminary data, assuming log_10_(GMT(PRNT_50_ at Day 0)) = log_10_(27) = 1.4 with a standard deviation of 0.9, a sample size of 80 individuals per group would provide 80% power to detect a difference in log_10_ GMT of 0.4 or greater at the 5% significance level. The same calculation applies in each stratum. We aimed for at least 100 individuals in each study arm to allow for the possibility of withdrawal from the study prior to the day 28 assessment.

### Laboratory analysis

Blood samples were delivered to our study laboratory at the University of Hong Kong as soon as possible, with the optimal delivery time within 24h. Clotted blood samples were stored in a refrigerated container at 2-8°C immediately after collection and while in transit to the central laboratory. Sera were extracted from the clotted blood samples within 48 hours after collection, divided into 2-4 aliquots, and stored at -80°C until subsequent serologic testing. Heparinized blood samples were kept at room temperature upon collection and while in transit and processed within 24 hours of collection. Peripheral blood mononuclear cells (PBMCs) were isolated from the heparinized (whole) blood using Ficoll-Paque and leucosep tubes, divided into 2 aliquots, and cryopreserved in liquid nitrogen until further analysis for cell-mediated immune responses.

Details of serologic testing, including our in-house enzyme-linked immunosorbent assay (ELISA) for the receptor binding domain (RBD) of the spike protein, a surrogate virus neutralisation test (sVNT) (GenScript) and a plaque reduction neutralisation test (PRNT), have been described in our earlier study on the immunogenicity of third-dose BNT162b2 after two doses of CoronaVac^8^. We aimed to test sera collected on Day 0 and 28 with ELISA against ancestral virus and sVNT against both ancestral virus and Omicron variant in all participants, and with PRNT against both ancestral virus and Omicron variant in a randomly selected subset of 20 participants in each study arm. We have demonstrated a good correlation (r=0.77) between PRNT_50_ and sVNT neutralization percentage for ancestral virus in our earlier studies^14^. The ELISA has a dynamic range of between 0 to 5, although it was not designed as a quantitative assay. For ELISA a single 1:100 serum dilution and for sVNT a single 1:10 serum dilution was tested respectively. For PRNT, a serial two-fold serum dilutions from 1:10 to 1:320 were tested initially, and in participants with an initial PRNT_50_ titer of ≥320, additional PRNT using a serial two-fold serum dilutions from 1:40 to 1:1280 were performed. PRNT assays were carried out using ancestral SARS-CoV-2 BetaCoV/Hong Kong/VM20001061/2020 (GISAID EPI_ISL_412028) isolated in Vero-E6 cells (ATCC CRL-1586), the Pango lineage B.1.1.529 Omicron BA.1 subvariant designated hCoV-19/Hong Kong/VM21044713_WHP5047-S5/2021 (GISAID EPI_ISL_6716902) and BA.2 subvariant hCoV-19/Hong Kong/VM22000135_HKUVOC0588P2/2022 (GISAID EPI_ISL_9570707) isolated in Vero-E6 TMRSS2 cells (Vero E6 cells overexpressing TMPRSS2, kindly provided by Dr S Matsuyama and colleagues),^15^ and the passage level 3 virus aliquots were used. Cells were maintained in DMEM medium supplemented with 10% FBS and 100 U/ml penicillin–streptomycin (all from ThermoFisher Scientific, Waltham, MA, USA). The PRNT_50_, PRNT_80_ and PRNT_90_ titers were the highest serum dilutions neutralising ≥50%, ≥80% and ≥90% of input plaques, respectively. The WHO control serum provided by the National Institute for Biological Standards and Control 20/136 gave PRNT_50_ titers of 320 and 320 against the ancestral virus and 20 and 40 against the Omicron variant in two titrations.

The cell-mediated immune responses to the third dose COVID-19 vaccination, proxy by the cytokine production of IFNγ (in both CD4^+^ and CD8^+^ T cells) or IL4 (in CD4^+^ T cells only) of T cells, were assessed in a randomly-selected subset of 20 participants in each study arm by intracellular cytokine staining (ICS). All testing was done in two experiments with each experiment including 10 participants randomly selected from each study arm, and time point samples collected from the same participant tested in parallel in the same experiment. Samples with a positive value after subtracting the respective value of the DMSO control were classified as having a positive response (“responder”) (value of DMSO control for CD4^+^ IFNγ^+^ T cells: 0.001%; CD8^+^ IFNγ^+^ T cells: 0.00017%; CD4^+^ IL4^+^ T cells: 0.000039%). Furthermore, CD4^+^ and CD8^+^ T cell memory phenotypes (CD45RA/CD27) and cytokine polyfunctional quality (TNF /IL2) were assessed by gating on IFNγ^+^ cells for samples which were classified as CD4^+^ IFNγ^+^ T cell or CD8^+^ IFNγ^+^ T cell responders^16^. Cryopreserved PBMCs collected on Day 0, 7 and 28 were thawed and re-stimulated with an overlapping peptide pool representing the SARS-CoV-2 structural proteins (spike, nucleocapsid, envelope and membrane) (300nM) or DMSO (1% in RPMI) in two independent experiments. Cells were stimulated for a total of 28 hours at 37°C. Golgi Plug (BD) containing Brefeldin A (1% in PBS), and Golgi Stop (BD) containing Monensin (0.67% in PBS) was added at 24 hours during stimulation for further 4-hour incubation. The amino acid sequence of the peptide pools was based on βCoV/Hong Kong/VM20001061/2020 strain (GISAID ID: EPI_ISL_412028). Cells were stained with Zombie-NIR (all antibodies from Biolegend and clone used) followed by anti-human CD3-PE/Dazzle 594 (UCHT1), CD4-BV605 (OKT4), CD8-AlexaFluor700 (SK1), CCR7-PerCP/Cy5.5 (G043H7), PD-1-BV421 (NAT105), CD45RA-APC (HI100), and a dump channel containing CD19-BV510 (HIB19), CD56-BV510 (5.1H11) and CD14-BV510 (M5E2).

Following cell permeabilization, intracellular staining with anti-IFNγ-FITC (4S.B3), IL4-PE (MP4-25D2) and TNFα-BV711 (Mab11), IL2-PECy7 (MQ1-17H12), was carried out before acquisition of samples. Stained cells were acquired via flow cytometry (AttuneNxT) and analysed by FlowJo v10. All samples met the cell viability cut-off of at least 30% and thus have been included in the analyses.

### Statistical analysis

We included all available data on participants who received a third dose of vaccination in our trial. For post-vaccination reactions, solicited local and systemic events or symptoms were presented as frequency (%) among participants who reported health status for ≥7 days in the week following receipt of the third dose. We compared the proportions of the participants with adverse events between the two study arms in individuals who had a prior recipient of two-dose BNT162b2, or who had a prior recipient of two doses of CoronaVac, using the Chi-squared test or Fisher’s exact test. Severity score as a proxy of interference to usual activities was calculated by assigning 0-reactions absent, 1-mild, 2-moderate and 3-severe and taking the average over the week after vaccination, and compared between groups using Student’s t-tests.

For antibody responses, for sVNT measured negative neutralization percentages were transformed to zero. PRNT titers were taken as the reciprocal of the serum dilution and were interval-censored, e.g. a sample that was able to neutralise virus at a 1:20 dilution but not at a 1:40 dilution was reported as 20 to indicate that the titer was ≥20 and <40. We estimated the arithmetic mean of SARS-CoV-2 surrogate virus neutralization percentages and concentrations of SARS-CoV-2 Spike RBD IgG (proxy by OD_450_) on Day 28. For PRNT titers, we imputed titers <10 with the value 5 and titers ≥1280 with the value 2560, and then transformed to log_2_ scale for the estimation of GMT and MFR, and significant testing. We compared antibody levels between Day 0 and Day 28 using Wilcoxon signed rank tests. We compared antibody levels at Day 28 between study arms using Mann-Whitney U tests. For cell-mediated response, we transformed CD4^+^ IFNγ^+^ T cell, CD8^+^ IFNγ^+^ T cell and CD4^+^ IL4^+^ T cell responses to log10 scale for the estimation of geometric means and fold-rises and significance testing. CD4^+^ and CD8^+^ T cell memory phenotypes and cytokine polyfunctional quality were assumed zeros for CD4^+^/CD8^+^ IFNγ^+^ T cell non-responders. We compared T cell responses between Day 0 and Day 7, and between Day 0 and Day 28, using Wilcoxon signed rank tests. We compared T cell responses at Day 7, and at Day 28, between study arms using Mann-Whitney U tests.

We compared the cumulative incidence of infections in each study arm using Kaplan-Meier curves and compared the incidence of infections using proportional hazards model. The proportional hazards model was specified on a calendar time axis with follow up starting from the start of Hong Kong’s fifth wave on 1 January 2022 or on the date of third dose if later, and ending at the earliest of the time of infection, the time of receiving a fourth dose, the time of withdrawing from the study, or the censoring date on 31 May 2022. If a participant reported more than one instance of positive result by RAT or PCR, the earliest date of identification was considered the time of infection. All statistical analyses were conducted using R version 4.2.1 (R Foundation for Statistical Computing, Vienna, Austria).

## RESULTS

From 12 November 2021 through 27 January 2022, we screened 994 individuals and 818 (82%) passed the initial eligibility assessment (**Figure 1, Figure S1**). We randomized 364 individuals to the CC-C (178) and CC-B (186) arms, and 406 individuals to the BB-C (202) and BB-B (204) arms. After final confirmation of eligibility, we collected baseline (Day 0) blood samples from 451 participants and then administered third doses, including 101 (57%) CC-C participants, 118 (63%) CC-B participants, 118 (58%) BB-C participants, and 114 (56%) BB-B participants. Participant characteristics were generally comparable between the CC-C and CC-B arms, and between the BB-C and BB-B arms (**Table 1**). Participants in the CC-C arm (median aged 58 years, IQR 50-64) were slightly older than those in CC-B (52 years, 43-58) (p = 0.01). For all 451 vaccinated participants, most reported receiving the second dose 6-8 months earlier (**Table 1**). A subset of 156 (35%) participants agreed to provide PBMCs samples on Day 0, 7 and Day 28 to evaluate cell-mediated immune responses (“CMI group”).

**Table 1.**
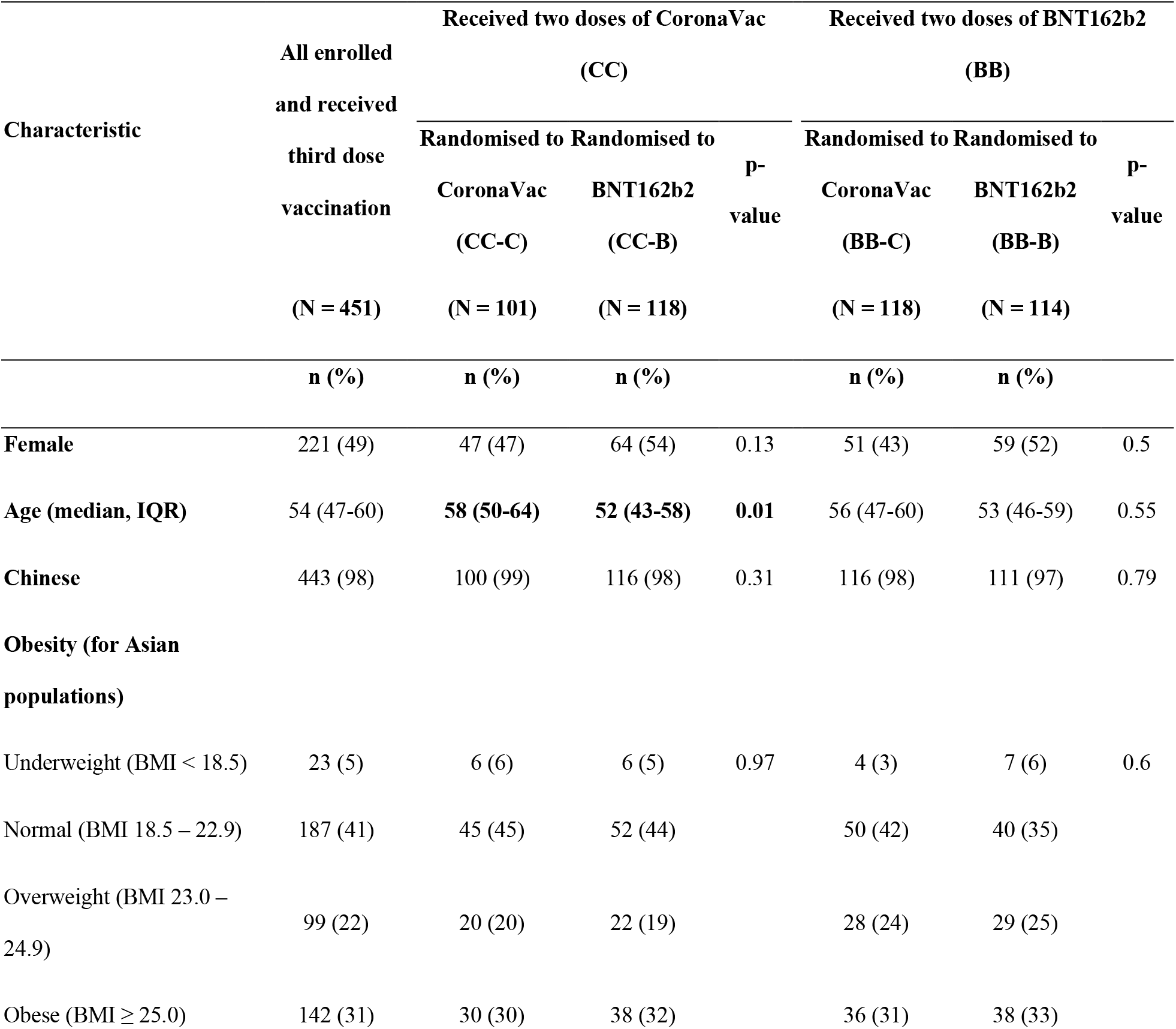
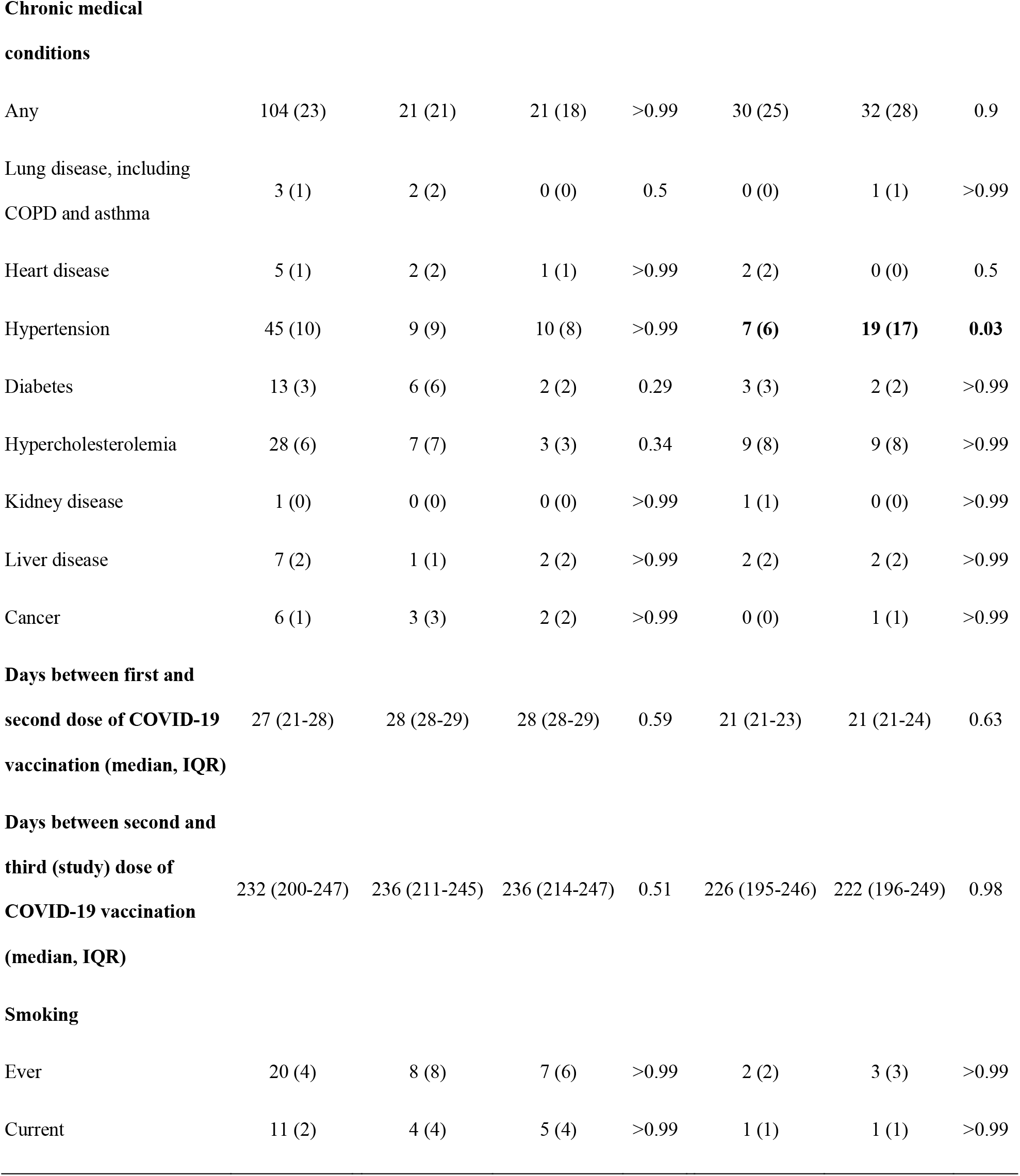
Characteristics of all vaccinated participants at baseline. We enrolled adults who previously received two doses of CoronaVac or BNT162b2, and randomly assigned participants to receive either vaccine as a third dose, i.e. the four study arms included participants who previously received two-dose CoronaVac and were randomised to receive a homologous third dose of CoronaVac (“CC-C”) or a heterologous third dose of BNT162b2 (“CC-B”), and participants who previously received two-dose BNT162b2 and were randomised to receive a heterologous third dose of CoronaVac (“BB-C”) or a homologous third dose of BNT162b2 (“BB-B”). Differences with p-values ≤ 0.05 were highlighted in bold.

**Figure 1.**
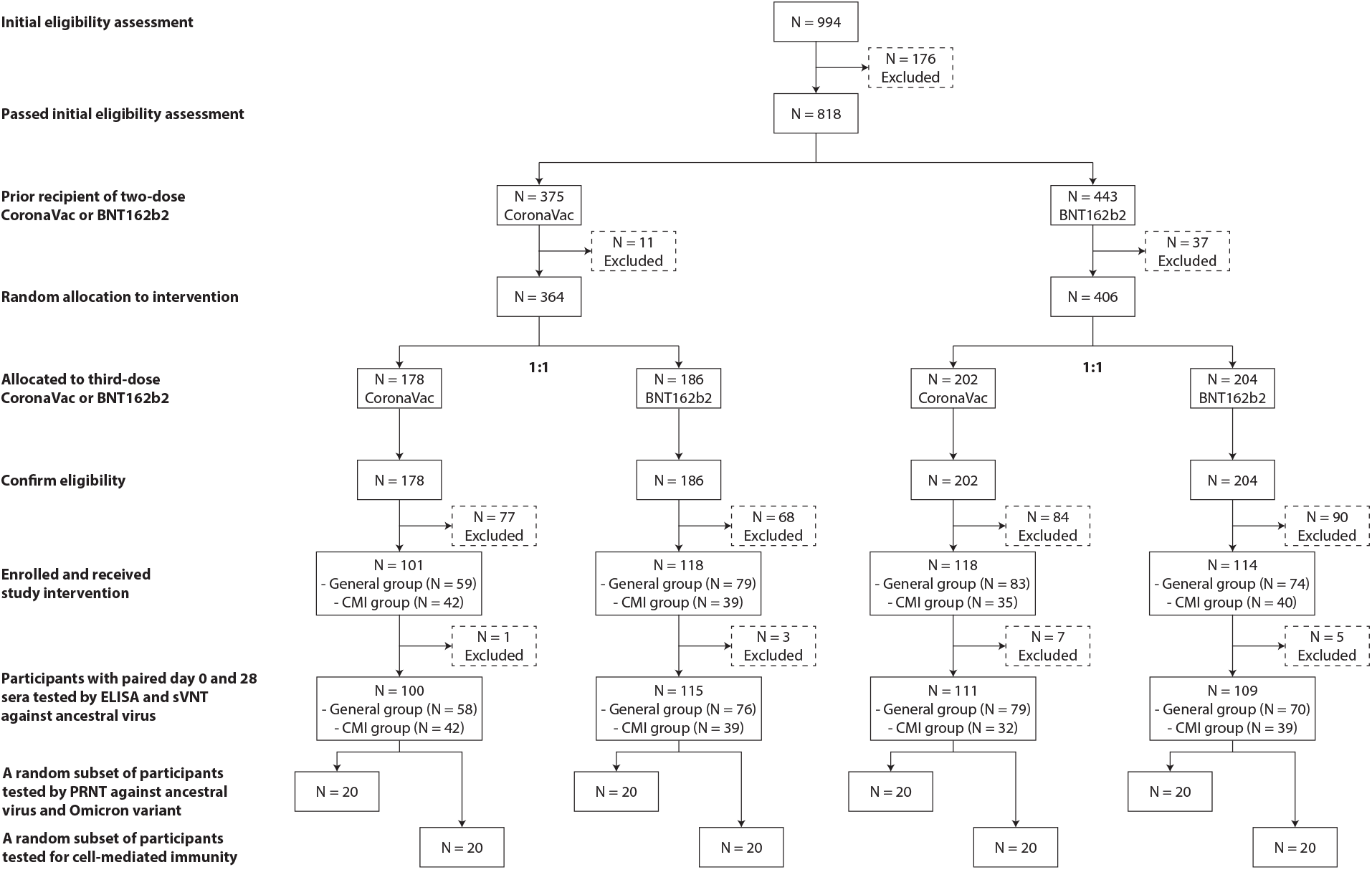
Flow chart of participant enrolment for this open-label randomised trial of CoronaVac or BNT162b2 provided as a third vaccine dose in adults who previously received two doses of either vaccine. A more detailed flow chart with reasons for exclusion at each stage is provided in Supplementary Figure S1.

We collected information on post-vaccination reactions in 424 (94%) participants, including 193 (64%) who reported ever feeling unwell after vaccination, with significantly more frequent reactions in recipients of a third dose of BNT162b2 that mostly subsided within 7 days (**Figure 2, Table S3**). Fever ≥38.0ºC was reported by about 10% of participants in both BB-B and CC-B arms, but by none in BB-C and CC-C arms (**Table S3**). Among 440 participants who reported information on medical care and hospitalisation, 10 reported seeking ambulatory care within one month of the third dose, including 4 (1 in CC-C and 3 in CC-B) for discomfort associated with vaccination (**Table S4**). One BB-B participant reported hospitalisation within a month of vaccination for unrelated reasons (**Table S4**).

**Figure 2.**
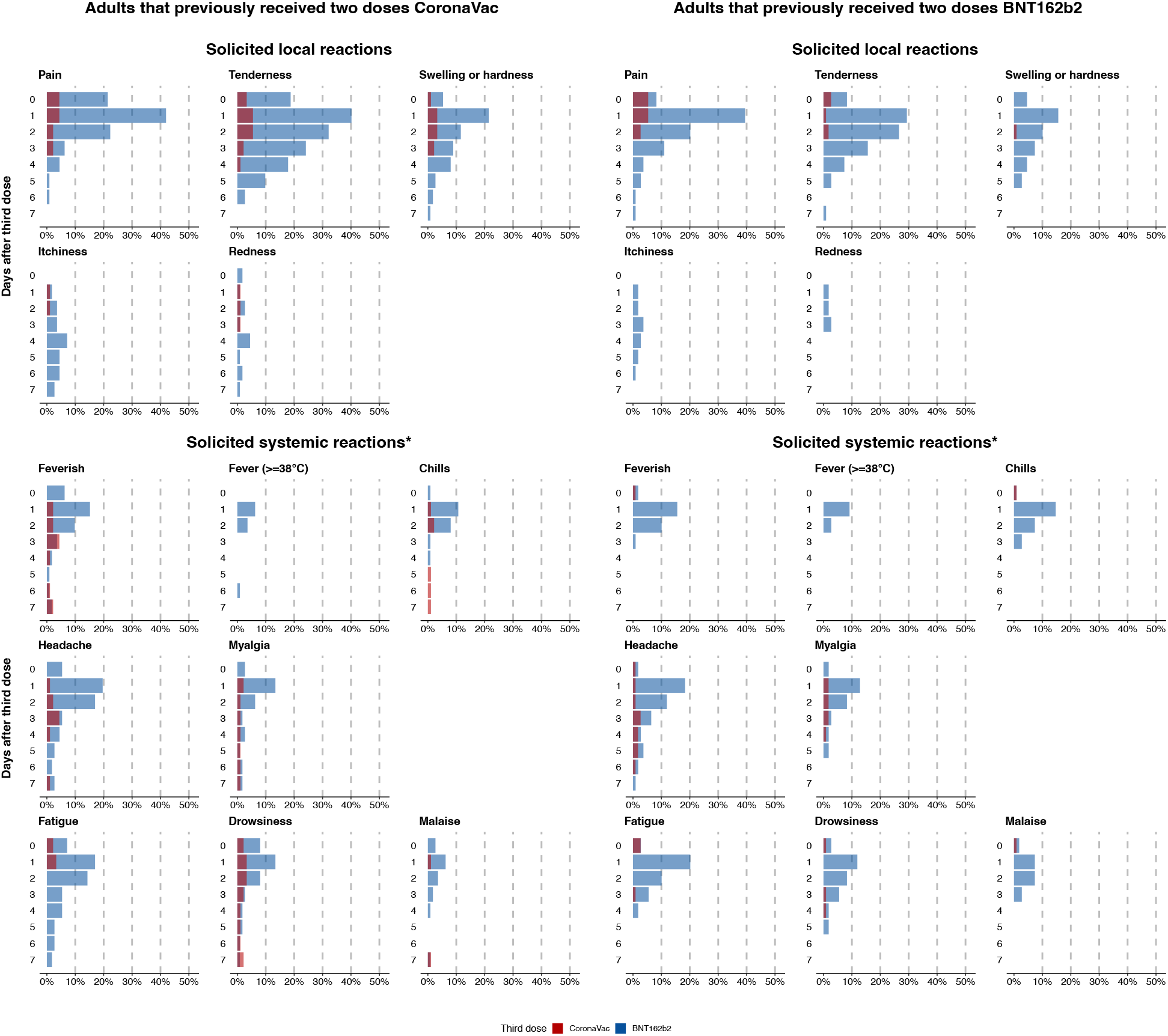
Solicited local and systemic reactions during the 7 days after a randomised third dose of CoronaVac or BNT162b2 vaccine in adults who previously received two doses of either vaccine. For solicited systemic reactions only the most frequently reported reactions are shown. The four study arms included participants who previously received two-dose CoronaVac and were randomised to receive a third dose of CoronaVac (“CC-C”) (red, left panel) or BNT162b2 (“CC-B”) (blue, left panel), and participants who previously received two-dose BNT162b2 and were randomised to receive a third dose of CoronaVac (“BB-C”) (red, right panel) or BNT162b2 (“BB-B”) (blue, right panel).

Third-dose vaccination significantly increased neutralising antibodies, measured as sVNT inhibition percentage or PRNT_50_ titers, against the ancestral virus, Omicron BA.1 and BA.2 on Day 28 from baseline in all four study arms regardless of third dose vaccine type (**Figure 3, Table S5**). In particular, third-dose vaccination increased PRNT_50_ titers against ancestral virus by 14-, 94-, 3- and 19-folds, and against Omicron BA.2 by 1-, 16-, 1- and 13-folds, in CC-C, CC-B, BB-C and BB-B arms respectively (**Figure S2A**). In both assays, antibody responses to a BNT162b2 third dose were substantially and statistically significantly greater than responses to a CoronaVac third dose regardless of prior two-dose vaccine type (**Figure 3, Figure S2A, Table S5**). Similar significant differences between study arms were observed when comparing binding antibodies against ancestral virus by ELISA (**Figure S3, Table S6**), or when comparing neutralizing antibodies evaluated at higher neutralization inhibition thresholds (PRNT_80_ or PRNT_90_ titers) (**Figure S4, Table S6**).

**Figure 3.**
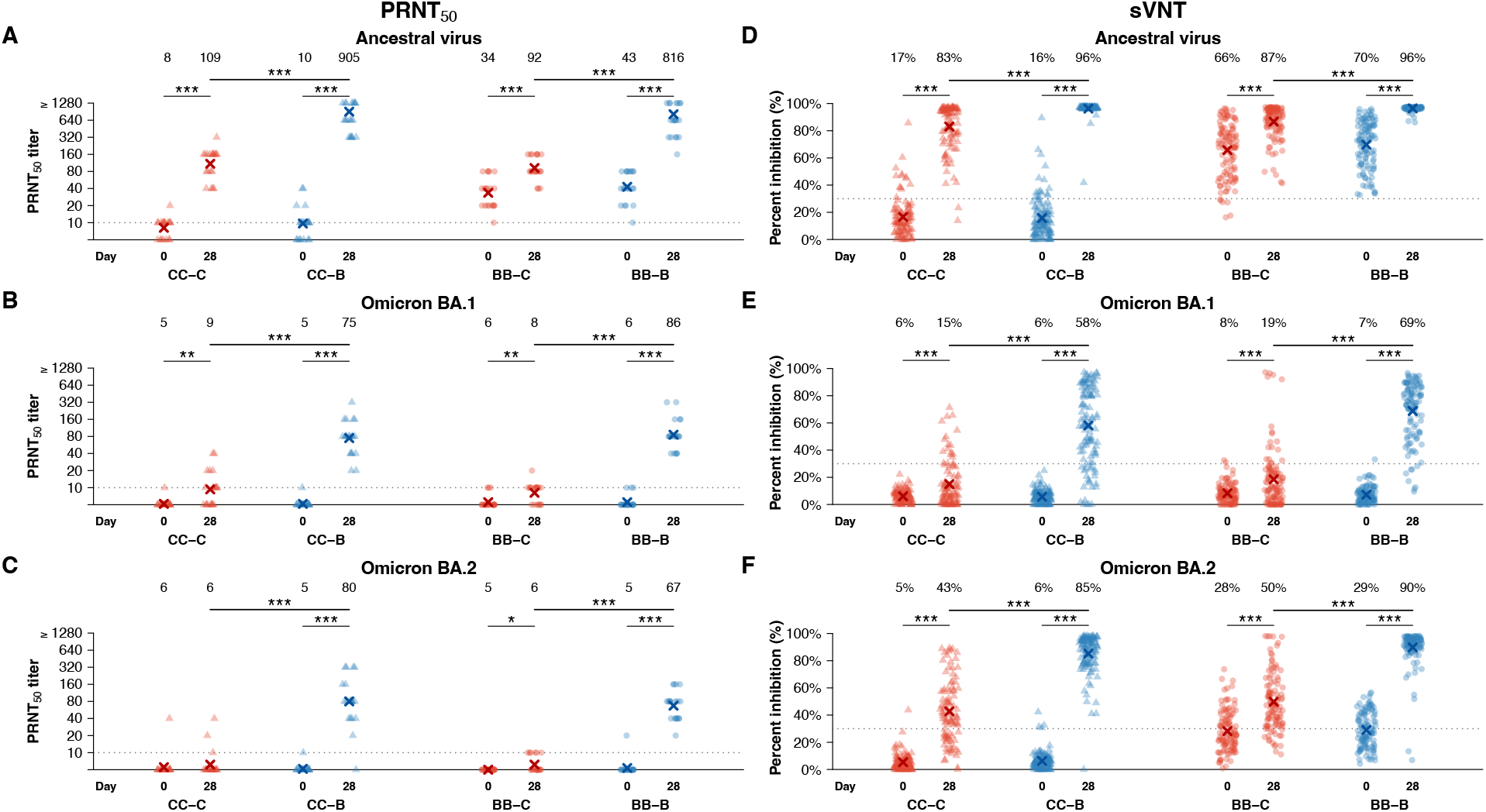
Serum neutralising antibodies measured by live virus plaque reduction neutralization test (PRNT) against (A) ancestral SARS-CoV-2 virus, (B) Omicron BA.1 and (C) Omicron BA.2 variants, or by (D-F) surrogate virus neutralization test (sVNT) respectively, at baseline and 28 days after a randomised third dose of CoronaVac or BNT162b2 vaccine in adults who previously received two doses of either vaccine. PRNT titers were evaluated with endpoint at 50% inhibition (PRNT_50_). Data on PRNT was available from a random subset of 20 participants selected from each study arm, while data on sVNT was available from all participants with paired sera. The four study arms included participants who previously received two-dose CoronaVac and were randomised to receive a third dose of CoronaVac (“CC-C”) or BNT162b2 (“CC-B”), and participants who previously received two-dose BNT162b2 and were randomised to receive a third dose of CoronaVac (“BB-C”) or BNT162b2 (“BB-B”). The symbol X and the numbers above each panel indicate the mean level. P-values ≤ 0.05 *, ≤ 0.01 **, ≤ 0.001 ***

In a subset of 20 participants randomly selected from each study arm (**Table S2**), we found that vaccination significantly increased the number of IFNγ^+^ CD4^+^ T cells (indicative of a Th1 response) in both CC-C and CC-B arms on Day 7, which remained significantly elevated from Day 0 on Day 28 (**Figure 4A, Figure S2B**). There was no significant change in IFNγ^+^ CD4^+^ T cells in both BB-C and BB-B arms on both Day 7 and Day 28 (**Figure 4A, Figure S2B**).

**Figure 4.**
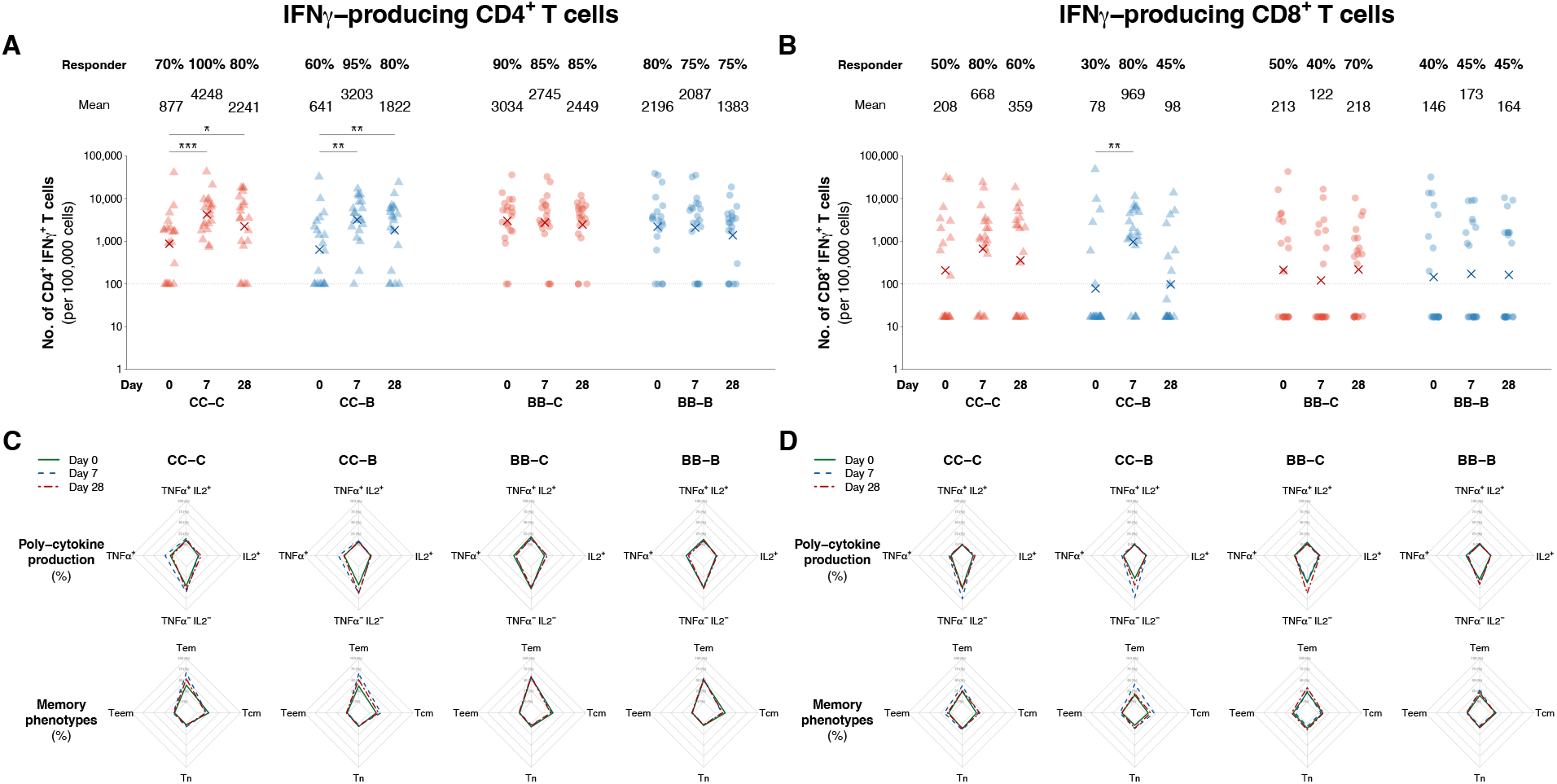
IFNγ-producing (A) CD4^+^ and (B) CD8^+^ T cell responses against structural peptides of ancestral SARS-CoV-2 virus, and (C-D) their poly-cytokine production (TNFα and IL-2) and memory phenotype respectively, at baseline, and 7 and 28 days after a randomised third dose of CoronaVac or BNT162b2 vaccine in adults who previously received two doses of either vaccine. Data was available from a random subset of 20 participants selected from each study arm. The four study arms included participants who previously received two-dose CoronaVac and were randomised to receive a third dose of CoronaVac (“CC-C”) or BNT162b2 (“CC-B”), and participants who previously received two-dose BNT162b2 and were randomised to receive a third dose of CoronaVac (“BB-C”) or BNT162b2 (“BB-B”). For **(A-B)**, the dotted lines represent the lower limits of detection based on DMSO controls and participants with values above these thresholds are considered as responders: 0.001% for CD4^+^ IFNγ^+^ T cells and 0.00017% for CD8^+^ IFNγ^+^ T cells. The numbers above each panel indicate the mean level, while the percentages indicate the proportion of responder. P-values ≤ 0.05 *, ≤ 0.01 **, ≤ 0.001 *** For **(C-D)**, the grey lines represent percentages of IFNγ-producing cells from 0% to 100% with increments of 25%, and the position of measurement on line represents the mean value of the 20 participants. Tn: naïve T cells, Tcm: central memory T cells, Tem: effector memory T cells, Teem: terminal effector memory T cells.

Separately, 30-50% participants in all arms had IFNγ^+^ CD8^+^ T cells (indicative of a CTL response) at baseline, but these were only significantly boosted in CC-B arm on Day 7 and contracted by Day 28 (**Figure 4B, Figure S2B**). Meanwhile, IL4^+^ CD4^+^ T cells (indicative of a Th2 response) were not significantly boosted by third doses in any arm (**Figure S2B, Figure S5**). There were no significant differences in the magnitude of CD4^+^ and CD8^+^ T cell responses between third doses of CoronaVac or BNT162b2 (**Table S6, Table S7**). We also evaluated the multi-cytokine production (TNFα and IL-2) and memory phenotype of IFNγ-producing CD4^+^ or CD8^+^ T cells. We observed a bias towards single cytokine responses (IFNγ^+^) as the majority of the T cell responses, but expansions upon vaccination of double cytokine responses (IFNγ^+^ with TNFα^+^ or IL-2^+^) in CC-C and CC-B; and T effector memory responses predominated across all arms and were further expanded in CC-C and CC-B (**Figure 4C and 4D, Table S5**).

We identified 58 SARS-CoV-2 infections by 31 May 2022, within 4-6 months of receipt of the third dose of vaccination. During this period the Omicron BA.2 variant was locally circulating. The number of infections identified by arm was 13/85 (15.3%) in CC-C, 16/104 (15.4%) in CC-B, 16/96 (16.7%) in BB-C and 13/93 (14.0%) in BB-B arms, with no statistically significant difference detected between the CC-C and CC-B arms (p = 0.93) nor between the BB-C and BB-B arms (p = 0.56) (**Figure 5**). Comparing BB-B and BB-C groups, the hazard ratio for a BNT162b2 third dose compared to a CoronaVac third dose was 0.80 (95% confidence interval: 0.39, 1.67) corresponding to a relative vaccine efficacy of 20%, while the hazard ratio for CC-B versus CC-C was 0.97 (95% confidence interval: 0.47, 2.01) corresponding to a relative vaccine efficacy of 3%. Additional information on active surveillance is available in **Supplementary Materials**.

**Figure 5.**
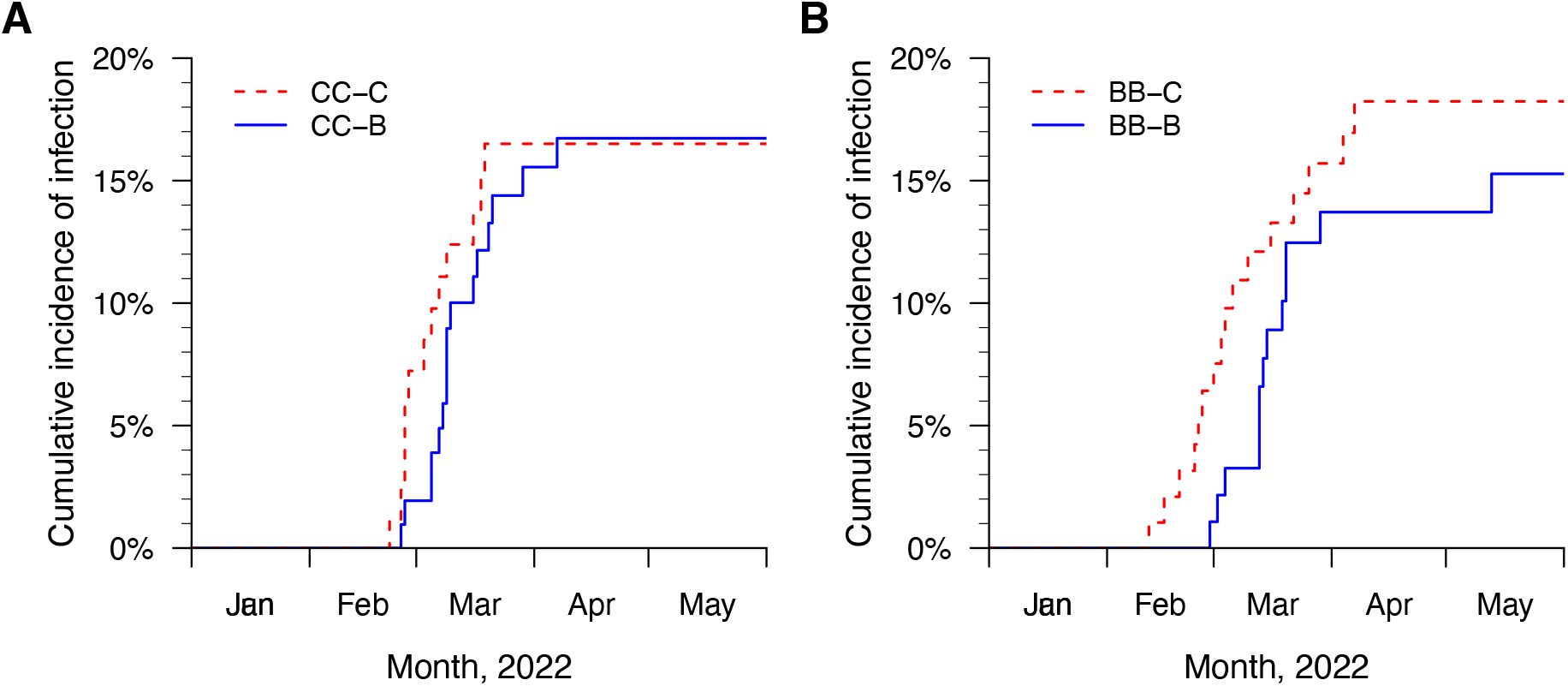
Cumulative incidence of SARS-CoV-2 infection after a randomised third dose of CoronaVac or BNT162b2 vaccine, in adults who previously received two doses of (A) CoronaVac or (B) BNT162b2, from after third-dose vaccination until 31 May 2022 and during the period when the Omicron BA.2 subvariant was circulating. SARS-CoV-2 infections were identified by rapid antigen test or PCR. The four study arms included participants who previously received two-dose CoronaVac and were randomised to receive a third dose of CoronaVac (“CC-C”) or BNT162b2 (“CC-B”), and participants who previously received two-dose BNT162b2 and were randomised to receive a third dose of CoronaVac (“BB-C”) or BNT162b2 (“BB-B”).

## DISCUSSION

Here, we have conducted a comprehensive assessment of the reactogenicity, antibody response, T cell responses and risk of infection in a randomised trial of homologous and heterologous boosting with BNT162b2 and CoronaVac. Our population had minimal COVID-19 infection history prior to all three doses of vaccination,^17^ and absence of known prior COVID-19 infection was one of the inclusion criteria. Furthermore, SARS-CoV-2 infection attack rate in the population of Hong Kong assessed by population based sero-surveillance studies was approximately 1% prior to the fifth wave of infections (unpublished data), thus likelihood of unsuspected infection in this cohort prior to vaccination was low. When evaluating the ability to boost immune response by a third vaccine dose, we showed that regardless of the combinations, a third vaccine dose increased neutralising antibody against all ancestral virus, the Omicron BA.1 and BA.2 variants significantly from baseline (**Figure 3**). The phenotype of the T cell responses was predominantly effector memory and single cytokine producing across all vaccine arms, but CD4^+^ and CD8^+^ T cell responses were only boosted in CC-C and CC-B, and no differences were found between all arms in total magnitude of T cell responses, phenotype or proportion responders (**Figure 4**). Prior influenza research has suggested the differentiation state of T cell responses can impact recall potential at infection and protection from disease.^18^ Thus, mRNA vaccine as a booster substantially increased antibody response, and those primed with two-dose inactivated vaccines by receiving mRNA or inactivated vaccine booster will benefit the T cell response, each providing an immunological layer against COVID-19. These findings suggest there is immune benefit of administering either a homologous or heterologous third dose six months after the second dose, especially more prominent in adults who initially received two doses of an inactivated vaccine.

The humoral and cellular immune results taken together would be indicative of superior efficacy of a third dose of BNT162b2 but that is not what we observed (**Figure 5**). Rates of infection, with Omicron BA.2 predominant in the community, were very similar among study arms (**Figure 5**). This discrepancy may indicate the challenge of using neutralising antibody as a correlate or mediator of protection against infection,^19,20^ at least in comparisons between vaccination strategies of mRNA and inactivated vaccines. Although neutralizing antibody did appear to be a correlate of protection when comparing RNA and adenovirus vectored vaccines and in comparisons between vaccinated and unvaccinated individuals,^21-23^ antibody alone may not explain the protection provided by inactivated vaccines, especially against viruses such as Omicron which have high escape from neutralizing antibody elicited by ancestral virus. T-cell responses may also play a role, as well as potentially non-neutralizing antibodies to internal virus genes such as the nucleocapsid. A discordance between neutralizing antibody titres and vaccine effectiveness against severe disease was also noted by us previously.^24^ Such differences between vaccine types may not always have been accounted for in immunobridging studies^25^.

The CD4^+^ helper T cell response, which has been shown to confer heterologous protection against lung damage from infection by Beta variant in mice,^26^ was significantly boosted and maintained by both homologous (CC-C) and heterologous (CC-B) third dose after two doses of inactivated vaccine (**Figure 4**). This is consistent with earlier findings,^27,28^ and these studies additionally suggested cross-reactivity against the Omicron variant by a third dose. Conversely, there was no boost of T cell responses neither by a homologous nor heterologous third dose after two doses of BNT162b2 (**Figure 4**), however a high level of responders at baseline was observed in both arms (**Figure 4A and 4B**). The CD8^+^ cytotoxic T cell response, which may have a role in limiting COVID-19 disease severity,^29,30^ was significantly boosted at Day 7 although not maintained to memory (Day 28) by heterologous third-dose BNT162b2 after two doses of CoronaVac (**Figure 4**). While the initial two doses were not randomized and participants did differ in a number of ways (**Table S9**), more participants who previously received two-dose BNT162b2 (80-90% responders) had CD4^+^ T cell responses compared to those who previously received two-dose CoronaVac (60-70% responders), and similar proportions of participants had CD8^+^ T cell response at baseline between all arms (30-50% responders, **Figure 4**). In addition, the baseline CD4^+^ T cell response of participants who received prior two doses of BNT162b2 were similar to the levels after third-dose vaccination in participants received prior two doses of CoronaVac (**Figure 4**). These may suggest a ceiling effect on the T cell response among participants who previously received two doses of BNT162b2 and explain the lack of boost in cell-mediated immunity in the BB-C and BB-B arms, which has also been observed elsewhere.^28,31^

Our study has several limitations. First, due to logistical challenges, we were only able to randomise at the time vaccination appointment was made but not at vaccination, and nearly half of randomized individuals dropped out before vaccination. However, the rates of dropout were similar across study arms, and we have taken great care in the randomisation and allocation concealment so that individuals dropping out were not aware of their allocated vaccine type. Comparisons between study arms did not identify significant differences in most baseline characteristics measured, except vaccinated participants in CC-B were on average 6 years younger than those in CC-C, and vaccinated participants in BB-B were over twice more likely to have hypertension than those in BB-C (**Table 1**). We also did not observe any significant differences in all antibody and cell-mediated response measured at baseline (**Table S6**). Second, in our study we did not include an unvaccinated control group, nor a two-dose comparison group, to evaluate the additional benefits of third dose over existing two doses. Third, we have only studied neutralising antibodies and T cell response, while other branches of immunity such as non-neutralising antibodies^32,33^ may also contribute to protection and may explain the discrepancy in our immunologic and efficacy data. Finally, our study population consists of adults whom first immune priming was from either mRNA or inactivated vaccination, which is different to many parts of the world where the first immune priming was from natural infection. This might lead to differences in subsequent immune response,^34^ vaccine effectiveness^35^ and duration of protection,^36^ and our study findings should be interpreted with these considerations.

In conclusion, our results suggest there is immune benefit in both administering a homologous or heterologous third dose 6 months after two doses of inactivated or mRNA vaccination, with similar levels of protection against infection provided by all four combinations. Mass vaccination programmes that offer both inactivated and mRNA vaccines, with explanation on their potential advantages and disadvantages, will help individuals in deciding their preferred option, allow flexibility in vaccine deployment, and encourage vaccine uptake.^37^ Our finding that neutralizing antibody may not be the dominant correlate of protection for inactivated vaccines, especially against SARS-CoV-2 variants such as Omicron that have significant capacity to evade neutralizing antibody, needs to be considered in the development of “variant proof” COVID-19 vaccines or vaccines broadly protective against sarbecoviruses currently in development.^38^

## Data Availability

Data to reproduce the results shown here will be posted on github after peer review and publication of our article.

## Acknowledgements

We gratefully acknowledge colleagues including Teresa So, Justin Wan and Eileen Yu for technical support in preparing and conducting this study; Anson Ho for setting up the database; Julie Au and Lilly Wang for administrative support; Hetti Cheung, Victoria Wong, Bobo Yeung at HKU Health System; Cindy Man and other colleagues at the HKU Community Vaccination Centres at Gleneagles Hospital; and all the study participants for facilitating the study.

## Funding

This project was supported by the Health and Medical Research Fund (grant no. COVID19F09), and the Theme-based Research Scheme of the Research Grants Council of the Hong Kong Special Administrative Region, China (grant no. T11-705/21-N). BJC is supported by the National Institute of Allergy and Infectious Diseases, National Institutes of Health, Department of Health and Human Services, under contract no. 75N93021C00015, and a RGC Senior Research Fellow Scheme grant (HKU SRFS2021-7S03) from the Research Grants Council of the Hong Kong Special Administrative Region, China. The funding bodies had no role in the design of the study, the collection, analysis, and interpretation of data, or writing of the manuscript.

## Author contributions

All authors meet the ICMJE criteria for authorship. Each author’s contributions to the paper are listed below according to the CrediT model:

Conceptualization: NHLL, GML, BJC.

Methodology: NHLL, SMSC, CAC, JSMP, SAV, BJC.

Formal analysis: NHLL, CAC, MM-S, SAV, BJC.

Investigation: CAC, NYMA, LLHL, LCHT, KKHK, SC, LWCF, AWLC, KCKC, JKCL, YYN, PK, JZJ.

Funding acquisition: NHLL, GML, BJC.

Project administration: NHLL, SMSC, JSMP, SAV, BJC.

Supervision: NHLL, SMSC, DKMI, LLMP, JSMP, SAV, BJC.

Writing – original draft: NHLL, CAC, MM-S, JSMP, SAV, BJC.

Writing – review & editing: NHLL, SMSC, CAC, MM-S, NYMA, LLHL, LCHT, KKHK, SC, LWCF, AWLC, KCKC, JKCL, YYN, PK, JZJ, DKMI, LLMP, GML, JSMP, SAV, BJC.

## Competing interests

BJC consults for AstraZeneca, Fosun Pharma, GlaxoSmithKline, Moderna, Pfizer, Roche and Sanofi Pasteur. BJC has received research funding from Fosun Pharma. The authors report no other potential conflicts of interest.

## Notes

### Clinical Trial

Clinicaltrials.gov NCT05057169

### Author Declarations

The study protocol (Clinicaltrials.gov NCT05057169) was approved by the Institutional Review Board of the University of Hong Kong (ref: UW 21-492).

### Summary of Updates

Fixed typos in the abstract and table 1

