## Supplementary Information for "Homologous and heterologous boosting with CoronaVac and BNT162b2: a randomized trial (the Cobovax study)"

**Supplementary Methods**

The flow of participant enrolment, i.e. various stages of eligibility assessment, randomization, vaccination and selection for laboratory testing is described in **Figure S1**. Invitations to participate were extended to community-dwelling adults in Hong Kong through mass promotion efforts including advertisements in newspapers, transportation and social media platforms, mass mailing to residential estates, and invitation to and referrals from members of existing cohorts. Interested individuals completed an online application form or called the study hotline. After completing a standardized screening form to preliminarily screen for eligibility and classification into two groups based on prior receipt of two doses of CoronaVac or BNT162b2, the study was explained in detail and participants expressing willingness to participate were randomized to receive either CoronaVac or BNT162b2 (as the third dose) in a 1:1 ratio and assigned a vaccination appointment accordingly. Shortly before or on arrival at the vaccination appointment, we explained the study again, confirmed eligibility and obtained written informed consent, before administering the study vaccination.

At the vaccination appointment (Day 0), all eligible participants were invited to a designated community vaccination centre, which runs under the government's mass vaccination program and offers COVID-19 vaccination to the general public. Under the supervision of the research team, participants were asked to complete a standardised enrolment questionnaire which included information including demographics, medication use, vaccination history, medical conditions and other health-related information such as smoking status, general functional and health status, and socioeconomic status. We collected a 20ml blood sample from the participants, and then unblinded nurses from the community vaccination centres administered the allocated third-dose vaccine intramuscularly. Participants were asked to return on Day 28 for a blood draw and follow-up assessment. Further follow-up blood draws are scheduled at 182 days and 365 days after vaccination. In a voluntary subset of approximately 35% of participants we collected additional 10ml whole blood samples at day 0, 7 and 28 for analysis of cell-mediated immune responses (CMI). At each encounter for a blood draw we offered a HKD100 (USD13) incentive in the form of a supermarket gift voucher.

After vaccination, participants were provided with vaccination cards and tympanic thermometers, observed for 15-30 minutes for immediate events, and then provided with an e-diary and requested to record daily on possible (delayed) adverse events or symptoms for at least 7 days, or until all adverse events disappeared. In the e-diary, participants were first asked whether they have felt unwell at any time during the past 24 hours. In those who felt unwell, they were asked for the presence of a list of provided (solicited) local and systemic adverse events, and any other (unsolicited) symptoms. Unwell participants were also asked to rate whether the reported adverse events have interfered with usual activities (mild - symptoms easily tolerated and did not interfere usual activities; moderate - symptoms interfered but did not prevent usual activities; severe - symptoms were severe and prevented usual activities), on the use of over-the-counter medication for symptom relief, absent from work or school, seeking medical care and hospitalisation. Participants were also asked for any medical care including ambulatory care and hospitalisation after the third-dose vaccination on Day 28, 182 and 365.

From March 2022, all participants were invited to participate in active surveillance to identify COVID-19 and influenza virus infection. Before initiation, consented participants were first asked about whether they had done any rapid antigen test (RAT) or virologic (PCR) test for COVID-19 or influenza before, and if yes whether there was any past COVID-19 or influenza virus infection defined as a positive test result identified by a RAT or virologic test. Then during active surveillance, all consented participants were provided with complementary COVID-19 RAT kits and asked to test themselves every 4 days ("systematic monitoring"). For each COVID-19 RAT, participants were asked to report and upload a photo of the test result to the study team through an online questionnaire whether the test result was positive or not. Participants were also asked to report any other RAT or virologic (PCR) test results for COVID-19 or influenza, such as those identified via mandatory mass testing requested by the government or voluntary testing for work purposes; or if any symptoms appear. When a positive test result for COVID-19 or influenza was identified, or any symptoms had appeared regardless of the RAT result, participants were asked to complete an online symptom diary daily, to monitor disease severity, medication use and medical attention ("illness episode"). Starting from 4 days after the initiation of the symptom diary, participants were also asked to performed COVID-19 RAT daily until obtaining negative result for two consecutive days. Participants completed the symptom diary daily for at least 7 days, or until all symptoms disappeared with RAT negative for two consecutive days, whichever the later. All COVID-19 RAT positive result identified during systematic monitoring and illness episodes were verified with the uploaded photo whenever available. The active surveillance is continuing with information collected up to May 31 included in the present analyses.

### Supplementary Results on active surveillance of SARS-CoV-2 infection

378 (84%) participants agreed to participate in active surveillance, had reported information on prior COVID-19 infection, and participated in systematic monitoring through self-administered COVID-19 RAT at least once during the active surveillance period, including 85/101 (84%) in CC-C, 104/118 (88%) in CC-B, 96/118 (81%) in BB-C and 93/114 (82%) in BB-B arms. After receiving the third dose vaccination and before the start of active surveillance, 42 SARS-CoV-2 infections were identified either by RAT or PCR, including 10, 11, 11 and 10 participants in each arm respectively. During active surveillance from 11 March to 31 May 2022, all participants on average reported RAT result every 4.5 days for a median of 68.5 days (IQR 60, 76). 24 SARS-CoV-2 infections were identified either by systematic RAT or self-reported PCR, including 3, 8, 5 and 8 participants in each arm respectively. Daily symptom diary was triggered by a positive RAT or PCR result, or symptomatic illness, for 53 illness episodes from 52 participants, from which 22 SARS-CoV-2 infections were identified by RAT or PCR, including 3, 9, 3 and 7 participants in each arm respectively. Together, 58 SARS-CoV-2 infections were identified within 4-6 months after third-dose vaccination during which Omicron BA.2 variant was locally circulating, including 13/85 (15.3%) in the CC-C, 16/104 (15.4%) in the CC-B, 16/96 (16.7%) in the BB-C and 13/93 (14.0%) in the BB-B arms, with no significant difference detected between the CC-C and CC-B arms (p = 0.93) nor between the BB-C and BB-B arms (p = 0.56) (**Figure 5**). A list of all positive COVID-19 results, method of identification and verification can be found in **Table S8**.

### Supplementary Tables

**Table S1. Characteristics of 435 trial participants at baseline, whom paired Day 0 and 28 sera were tested by ELISA and surrogate virus neutralization test (sVNT) to evaluate antibodies against ancestral virus after third-dose vaccination.** 435 trial participants who provided paired Day 0 and 28 sera, and whom Day 28 sera were provided within 60 days after third-dose vaccination were included, i.e. the four study arms included participants who previously received two-dose CoronaVac and were randomised to receive a homologous third dose of CoronaVac ("CC-C") or a heterologous third dose of BNT162b2 ("CC-B"), and participants who previously received two-dose BNT162b2 and were randomised to receive a heterologous third dose of CoronaVac ("BB-C") or a homologous third dose of BNT162b2 ("BB-B"). Differences with p-values ≤ 0.05 were highlighted in bold. ELISA: enzyme-linked immunosorbent assay; sVNT: surrogate virus neutralisation test

| | | **All enrolled and received third dose vaccination** | **Received primary series of CoronaVac (CC)** | | | | **Received primary series of BNT162b2 (BB)** | | |
|---|---|---|---|---|---|---|---|---|---|
| | | | **Randomised to CoronaVac (CC-C)** | **Randomised to BNT162b2 (CC-B)** | ***p*** | | **Randomised to CoronaVac (BB-C)** | **Randomised to BNT162b2 (BB-B)** | ***p*** |
| | | **(n = 451)** | **(n = 101)** | **(n = 118)** | | | **(n = 118)** | **(n = 114)** | |
| | | **n (%)** | **n (%)** | **n (%)** | | | **n (%)** | **n (%)** | |
| **Female** | | 221 (49) | 47 (47) | 64 (54) | 0.13 | | 51 (43) | 59 (52) | 0.5 |
| **Age (median, IQR)** | | 54 (47-60) | **58 (50-64)** | **52 (43-58)** | **0.01** | | 56 (47-60) | 53 (46-59) | 0.55 |
| **Age group (years)** | | | | | | | | | |
| <18 | | 0 (0) | 0 (0) | 0 (0) | | | 0 (0) | 0 (0) | |
| 18 - 29 | | 22 (5) | 5 (5) | 5 (4) | | | 5 (4) | 7 (6) | |
| 30 - 39 | | 38 (8) | 8 (8) | 13 (11) | | | 8 (7) | 9 (8) | |
| 40 - 49 | | 100 (22) | 13 (13) | 34 (29) | | | 27 (23) | 26 (23) | |
| 50 - 59 | | 173 (38) | 36 (36) | 45 (38) | | | 47 (40) | 45 (39) | |
| 60 - 69 | | 99 (22) | 33 (33) | 20 (17) | | | 28 (24) | 18 (16) | |
| 70 - 79 | | 19 (4) | 6 (6) | 1 (1) | | | 3 (3) | 9 (8) | |
| ≥80 | | 0 (0) | 0 (0) | 0 (0) | | | 0 (0) | 0 (0) | |
| **Chinese** | | 443 (98) | 100 (99) | 116 (98) | 0.31 | | 116 (98) | 111 (97) | 0.79 |
| **Obesity** | | | | | | | | | |
| Underweight | | 23 (5) | 6 (6) | 6 (5) | 0.97 | | 4 (3) | 7 (6) | 0.6 |
| Normal | | 187 (41) | 45 (45) | 52 (44) | | | 50 (42) | 40 (35) | |
| Overweight | | 99 (22) | 20 (20) | 22 (19) | | | 28 (24) | 29 (25) | |
| Obese | | 142 (31) | 30 (30) | 38 (32) | | | 36 (31) | 38 (33) | |
| **Chronic medical conditions** | | | | | | | | | |
| Any | | 104 (23) | 21 (21) | 21 (18) | >0.99 | | 30 (25) | 32 (28) | 0.9 |
| Lung disease, including COPD and asthma | | 3 (1) | 2 (2) | 0 (0) | 0.5 | | 0 (0) | 1 (1) | >0.99 |
| Heart disease | | 5 (1) | 2 (2) | 1 (1) | >0.99 | | 2 (2) | 0 (0) | 0.5 |

| | | | | | | | | | |
|---|---|---|---|---|---|---|---|---|---|
| Hypertension | | 45 (10) | 9 (9) | 10 (8) | >0.99 | | **7 (6)** | **19 (17)** | **0.03** |
| Diabetes | | 13 (3) | 6 (6) | 2 (2) | 0.29 | | 3 (3) | 2 (2) | >0.99 |
| Hypercholesterolemia | | 28 (6) | 7 (7) | 3 (3) | 0.34 | | 9 (8) | 9 (8) | >0.99 |
| Kidney disease | | 1 (0) | 0 (0) | 0 (0) | >0.99 | | 1 (1) | 0 (0) | >0.99 |
| Liver disease | | 7 (2) | 1 (1) | 2 (2) | >0.99 | | 2 (2) | 2 (2) | >0.99 |
| Cancer | | 6 (1) | 3 (3) | 2 (2) | >0.99 | | 0 (0) | 1 (1) | >0.99 |
| **Days between first and second dose of COVID-19 vaccination (median, IQR)** | | 27 (21-28) | 28 (28-29) | 28 (28-29) | 0.59 | | 21 (21-23) | 21 (21-24) | 0.63 |
| **Days between second and third (study) dose of COVID-19 vaccination (median, IQR)** | | 232 (200-247) | 236 (211-245) | 236 (214-247) | 0.51 | | 226 (195-246) | 222 (196-249) | 0.98 |
| **Smoking** | | | | | | | | | |
| Ever | | 20 (4) | 8 (8) | 7 (6) | >0.99 | | 2 (2) | 3 (3) | >0.99 |
| Current | | 11 (2) | 4 (4) | 5 (4) | >0.99 | | 1 (1) | 1 (1) | >0.99 |

**Table S2. Characteristics of randomly selected trial participants at baseline, whom paired Day 0 and 28 sera were tested by plaque reduction neutralization test (PRNT) to evaluate neutralising antibodies against ancestral virus and Omicron BA.1 and BA.2 variants after third-dose vaccination, or whom Day 0, 7 and 28 PBMCs were tested by intracellular cytokine staining (ICS) to evaluate T cell response against structural peptides of ancestral SARS-CoV-2 virus after third-dose vaccination.** Two independent sets of 80 trial participants, 20 selected from each study arm, were randomly selected for PRNT or ICS. The four study arms included participants who previously received two-dose CoronaVac and were randomised to receive a third dose of CoronaVac ("CC-C") or BNT162b2 ("CC-B"), and participants who previously received two-dose BNT162b2 and were randomised to receive a third dose of CoronaVac ("BB-C") or BNT162b2 ("BB-B"). Differences with p-values ≤ 0.05 were highlighted in bold. PRNT: plaque reduction neutralisation test; ICS: intracellular cytokine staining; PBMCs: peripheral blood mononuclear cells.

| | **Randomly selected for PRNT** | | | | | | **Randomly selected for ICS** | | | | | |
|---|---|---|---|---|---|---|---|---|---|---|---|---|
| | **Received two-dose CoronaVac (CC)** | | | **Received two-dose BNT162b2 (BB)** | | | **Received two-dose CoronaVac (CC)** | | | **Received two-dose BNT162b2 (BB)** | | |
| | **Randomised to CoronaVac (CC-C)** | **Randomised to BNT162b2 (CC-B)** | ***p*** | **Randomised to CoronaVac (BB-C)** | **Randomised to BNT162b2 (BB-B)** | ***p*** | **Randomised to CoronaVac (CC-C)** | **Randomised to BNT162b2 (CC-B)** | ***p*** | **Randomised to CoronaVac (BB-C)** | **Randomised to BNT162b2 (BB-B)** | ***p*** |
| | **(n = 20)** | **(n = 20)** | | **(n = 20)** | **(n = 20)** | | **(n = 20)** | **(n = 20)** | | **(n = 20)** | **(n = 20)** | |
| | **n (%)** | **n (%)** | | **n (%)** | **n (%)** | | **n (%)** | **n (%)** | | **n (%)** | **n (%)** | |
| **Female** | 8 (40) | 11 (55) | 0.65 | 8 (40) | 7 (35) | >0.99 | 9 (45) | 8 (40) | >0.99 | 6 (30) | 13 (65) | 0.17 |
| **Age (median, IQR)** | 62 (53-67) | 57 (51-60) | 0.18 | 56 (48-61) | 52 (39-58) | 0.38 | 58 (51-65) | 50 (43-61) | 0.07 | 58 (51-64) | 52 (43-59) | 0.08 |
| **Age group (years)** | - | - | | - | - | | - | - | | - | - | |
| <18 | 0 (0) | 0 (0) | 0.38 | 0 (0) | 0 (0) | 0.24 | 0 (0) | 0 (0) | 0.42 | 0 (0) | 0 (0) | 0.15 |
| 18 - 29 | 0 (0) | 0 (0) | | 1 (5) | 2 (10) | | 0 (0) | 0 (0) | | 1 (5) | 1 (5) | |
| 30 - 39 | 1 (5) | 1 (5) | | 1 (5) | 3 (15) | | 0 (0) | 3 (15) | | 0 (0) | 2 (10) | |
| 40 - 49 | 2 (10) | 4 (20) | | 4 (20) | 4 (20) | | 5 (25) | 7 (35) | | 2 (10) | 6 (30) | |
| 50 - 59 | 7 (35) | 10 (50) | | 9 (45) | 7 (35) | | 6 (30) | 4 (20) | | 9 (45) | 7 (35) | |
| 60 - 69 | 7 (35) | 5 (25) | | 5 (25) | 1 (5) | | 7 (35) | 5 (25) | | 8 (40) | 3 (15) | |
| 70 - 79 | 3 (15) | 0 (0) | | 0 (0) | 3 (15) | | 2 (10) | 1 (5) | | 0 (0) | 1 (5) | |
| ≥80 | 0 (0) | 0 (0) | | 0 (0) | 0 (0) | | 0 (0) | 0 (0) | | 0 (0) | 0 (0) | |

| | | | | | | | | | | | | | | | |
|---|---|---|---|---|---|---|---|---|---|---|---|---|---|---|---|
| **Chinese** | 19 (95) | 20 (100) | >0.99 | | 19 (95) | 19 (95) | >0.99 | | 20 (100) | 20 (100) | >0.99 | | 20 (100) | 20 (100) | >0.99 |
| **Obesity** | - | - | | | - | - | | | - | - | | | - | - | |
| Underweight | 0 (0) | 1 (5) | 0.46 | | 0 (0) | 1 (5) | >0.99 | | **1 (5)** | **1 (5)** | **<0.01** | | 1 (5) | 3 (15) | 0.71 |
| Normal | 10 (50) | 6 (30) | | | 9 (45) | 8 (40) | | | **14 (70)** | **4 (20)** | | | 8 (40) | 7 (35) | |
| Overweight | 4 (20) | 4 (20) | | | 7 (35) | 7 (35) | | | **2 (10)** | **2 (10)** | | | 5 (25) | 3 (15) | |
| Obese | 6 (30) | 9 (45) | | | 4 (20) | 4 (20) | | | **3 (15)** | **13 (65)** | | | 6 (30) | 7 (35) | |
| **Chronic medical conditions** | - | - | | | - | - | | | - | - | | | - | - | |
| Any | 2 (10) | 4 (20) | 0.69 | | 6 (30) | 4 (20) | 0.75 | | 5 (25) | 3 (15) | 0.73 | | 6 (30) | 5 (25) | >0.99 |
| Lung disease, including COPD and asthma | 0 (0) | 0 (0) | >0.99 | | 0 (0) | 0 (0) | >0.99 | | 2 (10) | 0 (0) | 0.5 | | 0 (0) | 0 (0) | >0.99 |
| Heart disease | 0 (0) | 0 (0) | >0.99 | | 0 (0) | 0 (0) | >0.99 | | 1 (5) | 0 (0) | >0.99 | | 1 (5) | 0 (0) | >0.99 |
| Hypertension | 1 (5) | 2 (10) | >0.99 | | 0 (0) | 3 (15) | 0.25 | | 2 (10) | 1 (5) | >0.99 | | 0 (0) | 4 (20) | 0.12 |
| Diabetes | 0 (0) | 0 (0) | >0.99 | | 0 (0) | 2 (10) | 0.5 | | 1 (5) | 0 (0) | >0.99 | | 0 (0) | 1 (5) | >0.99 |
| Hypercholesterolemia | 1 (5) | 1 (5) | >0.99 | | 1 (5) | 2 (10) | >0.99 | | 1 (5) | 0 (0) | >0.99 | | 2 (10) | 2 (10) | >0.99 |
| Kidney disease | 0 (0) | 0 (0) | >0.99 | | 1 (5) | 0 (0) | >0.99 | | 0 (0) | 0 (0) | >0.99 | | 1 (5) | 0 (0) | >0.99 |
| Liver disease | 0 (0) | 0 (0) | >0.99 | | 0 (0) | 1 (5) | >0.99 | | 0 (0) | 0 (0) | >0.99 | | 0 (0) | 0 (0) | >0.99 |
| Cancer | 1 (5) | 0 (0) | >0.99 | | 0 (0) | 0 (0) | >0.99 | | 0 (0) | 0 (0) | >0.99 | | 0 (0) | 0 (0) | >0.99 |
| **Days between first and second dose of COVID-19 vaccination (median, IQR)** | **28 (28-28)** | **28 (28-30)** | **0.04** | | 21 (21-22) | 22 (21-24) | 0.19 | | 28 (28-28) | 29 (28-30) | 0.17 | | 21 (21-21) | 21 (21-22) | 0.49 |
| **Days between second and third (study) dose of COVID-19 vaccination (median, IQR)** | 238 (228-244) | 236 (215-247) | 0.67 | | 229 (214-234) | 224 (196-234) | 0.67 | | 234 (222-239) | 230 (219-243) | 0.72 | | 232 (222-241) | 230 (194-250) | 0.61 |
| **Smoking** | - | - | | | - | - | | | - | - | | | - | - | |
| Ever | 1 (5) | 0 (0) | >0.99 | | 0 (0) | 2 (10) | 0.5 | | 0 (0) | 0 (0) | >0.99 | | 0 (0) | 1 (5) | >0.99 |
| Current | 1 (5) | 0 (0) | >0.99 | | 0 (0) | 0 (0) | >0.99 | | 0 (0) | 0 (0) | >0.99 | | 0 (0) | 1 (5) | >0.99 |

**Table S3. Solicited local and systemic reactions anytime during the 7 days after a randomised third dose of CoronaVac or BNT162b2 vaccine in adults who previously received two doses of either vaccine.** The four study arms included participants who previously received two-dose CoronaVac and were randomised to receive a third dose of CoronaVac ("CC-C") or BNT162b2 ("CC-B"), and participants who previously received two-dose BNT162b2 and were randomised to receive a third dose of CoronaVac ("BB-C") or BNT162b2 ("BB-B"). Differences with p-values ≤ 0.05 were highlighted in bold.

| | **Received primary series of CoronaVac (CC)** | | | | **Received primary series of BNT162b2 (BB)** | | |
|---|---|---|---|---|---|---|---|
| | **Randomised to CoronaVac (CC-C)** | **Randomised to BNT162b2 (CC-B)** | ***p*** | | **Randomised to CoronaVac (BB-C)** | **Randomised to BNT162b2 (BB-B)** | ***p*** |
| | **(n = 91)** | **(n = 112)** | | | **(n = 112)** | **(n = 109)** | |
| | **n (%)** | **n (%)** | | | **n (%)** | **n (%)** | |
| ***Reported ever feeling unwell*** | **24 (26%)** | **77 (69%)** | **<0.001** | | **25 (22%)** | **67 (61%)** | **<0.001** |
| ***Duration of adverse reactions (Mean, SD)*** | **2.2 (1.8)** | **3.6 (2.2)** | **0.0245** | | 2.2 (1.4) | 3.0 (1.6) | 0.145 |
| ***Severity score for interfering ususal activities (Mean, SD)*** | **0.08 (0.18)** | **0.40 (0.44)** | **<0.001** | | **0.07 (0.17)** | **0.31 (0.34)** | **<0.001** |
| ***Body temperature (Median, IQR)*** | - | 38.2 (38.0,38.4) | - | | - | 38.4 (38.3,38.6) | - |
| ***Solicited local reactions*** | | | | | | | |
| **Pain** | **8 (8.8%)** | **59 (53%)** | **<0.001** | | **10 (8.9%)** | **50 (46%)** | **<0.001** |
| **Tenderness** | **8 (8.8%)** | **57 (51%)** | **<0.001** | | **5 (4.5%)** | **43 (39%)** | **<0.001** |
| **Swelling or hardness** | **5 (5.5%)** | **30 (27%)** | **<0.001** | | **1 (0.9%)** | **21 (19%)** | **<0.001** |
| **Itchiness** | **2 (2.2%)** | **11 (9.8%)** | **0.027** | | **0 (0%)** | **9 (8.3%)** | **0.001** |
| **Redness** | **1 (1.1%)** | **9 (8.0%)** | **0.025** | | **0 (0%)** | **6 (5.5%)** | **0.013** |
| ***Solicited systemic reactions*** | | | | | | | |
| **Headache** | **5 (5.5%)** | **32 (29%)** | **<0.001** | | **7 (6.2%)** | **28 (26%)** | **<0.001** |
| **Fatigue** | **4 (4.4%)** | **32 (29%)** | **<0.001** | | **4 (3.6%)** | **24 (22%)** | **<0.001** |
| **Feverish** | **7 (7.7%)** | **28 (25%)** | **0.001** | | **1 (0.9%)** | **22 (20%)** | **<0.001** |
| **Fever (≥38ºC)** | **0 (0%)** | **12 (11%)** | **0.001** | | **0 (0%)** | **11 (10%)** | **<0.001** |
| **Chills** | **4 (4.4%)** | **18 (16%)** | **0.008** | | **1 (0.9%)** | **22 (20%)** | **<0.001** |
| **Drowsiness** | **5 (5.5%)** | **24 (21%)** | **0.001** | | **2 (1.8%)** | **21 (19%)** | **<0.001** |
| **Myalgia** | **3 (3.3%)** | **21 (19%)** | **<0.001** | | **4 (3.6%)** | **17 (16%)** | **0.002** |
| **Malaise** | **2 (2.2%)** | **13 (12%)** | **0.011** | | **1 (0.9%)** | **15 (14%)** | **<0.001** |
| Pain at the injection arm | **2 (2.2%)** | **10 (8.9%)** | **0.043** | | **1 (0.9%)** | **13 (12%)** | **<0.001** |
| Arthralgia | 3 (3.3%) | 8 (7.1%) | 0.4 | | **2 (1.8%)** | **13 (12%)** | **0.003** |
| Loss of appetite | **0 (0%)** | **9 (8.0%)** | **0.005** | | **2 (1.8%)** | **10 (9.2%)** | **0.015** |
| Abdominal distention | 0 (0%) | 4 (3.6%) | 0.13 | | **1 (0.9%)** | **8 (7.3%)** | **0.018** |
| Nasal congestion | 2 (2.2%) | 7 (6.2%) | 0.2 | | **0 (0%)** | **7 (6.4%)** | **0.006** |
| Chest discomfort | 1 (1.1%) | 7 (6.2%) | 0.077 | | 2 (1.8%) | 7 (6.4%) | 0.1 |
| Dizziness | 4 (4.4%) | 12 (11%) | 0.1 | | 2 (1.8%) | 7 (6.4%) | 0.1 |
| Runny nose | **3 (3.3%)** | **13 (12%)** | **0.029** | | 4 (3.6%) | 6 (5.5%) | 0.5 |
| Sneezing | 2 (2.2%) | 7 (6.2%) | 0.2 | | 1 (0.9%) | 6 (5.5%) | 0.063 |
| Phlegm | 4 (4.4%) | 4 (3.6%) | >0.9 | | 1 (0.9%) | 6 (5.5%) | 0.063 |
| Cough | **3 (3.3%)** | **12 (11%)** | **0.045** | | 1 (0.9%) | 5 (4.6%) | 0.12 |
| Sore throat | 3 (3.3%) | 10 (8.9%) | 0.1 | | 3 (2.7%) | 5 (4.6%) | 0.5 |
| Nausea | 1 (1.1%) | 5 (4.5%) | 0.2 | | 4 (3.6%) | 5 (4.6%) | 0.7 |
| Insomnia | 2 (2.2%) | 8 (7.1%) | 0.2 | | 1 (0.9%) | 4 (3.7%) | 0.2 |
| Diarrhoea | **0 (0%)** | **6 (5.4%)** | **0.034** | | 3 (2.7%) | 3 (2.8%) | >0.9 |
| Body itching | 2 (2.2%) | 6 (5.4%) | 0.3 | | 1 (0.9%) | 3 (2.8%) | 0.4 |
| Flushing of the face | 2 (2.2%) | 4 (3.6%) | 0.7 | | 0 (0%) | 3 (2.8%) | 0.12 |
| Chest pain | 1 (1.1%) | 2 (1.8%) | >0.9 | | 1 (0.9%) | 3 (2.8%) | 0.4 |

| | | | | | | | |
|---|---|---|---|---|---|---|---|
| Abdominal pain | 0 (0%) | 1 (0.9%) | >0.9 | | 2 (1.8%) | 3 (2.8%) | 0.7 |
| Palpitations | **0 (0%)** | **9 (8.0%)** | **0.005** | | 3 (2.7%) | 2 (1.8%) | >0.9 |
| Skin rash | **0 (0%)** | **6 (5.4%)** | **0.034** | | 0 (0%) | 2 (1.8%) | 0.2 |
| Constipation | 2 (2.2%) | 5 (4.5%) | 0.5 | | 1 (0.9%) | 2 (1.8%) | 0.6 |
| Muscle spasms | 0 (0%) | 0 (0%) | - | | 0 (0%) | 2 (1.8%) | 0.2 |
| Ageusia | 0 (0%) | 1 (0.9%) | >0.9 | | 0 (0%) | 1 (0.9%) | 0.5 |
| Enlarged lymph nodes | 0 (0%) | 0 (0%) | - | | 0 (0%) | 1 (0.9%) | 0.5 |
| Face swelling | 0 (0%) | 0 (0%) | - | | 0 (0%) | 1 (0.9%) | 0.5 |
| Arm or leg swelling | 0 (0%) | 0 (0%) | - | | 2 (1.8%) | 1 (0.9%) | >0.9 |
| Vomiting | 0 (0%) | 0 (0%) | - | | 2 (1.8%) | 1 (0.9%) | >0.9 |
| Facial drooping/ weakness | 0 (0%) | 0 (0%) | - | | 0 (0%) | 1 (0.9%) | 0.5 |
| Nosebleeds | 1 (1.1%) | 2 (1.8%) | >0.9 | | 0 (0%) | 0 (0%) | - |
| Dyspnea | 0 (0%) | 2 (1.8%) | 0.5 | | 0 (0%) | 0 (0%) | - |
| Confusion | 0 (0%) | 1 (0.9%) | >0.9 | | 0 (0%) | 0 (0%) | - |
| Anosmia | 0 (0%) | 0 (0%) | - | | 0 (0%) | 0 (0%) | - |
| Conjunctivitis | 0 (0%) | 0 (0%) | - | | 0 (0%) | 0 (0%) | |
| Loss of consciousness | 0 (0%) | 0 (0%) | - | | 0 (0%) | 0 (0%) | - |
| Seizure | 0 (0%) | 0 (0%) | - | | 0 (0%) | 0 (0%) | - |

**Table S4. Reasons for seeking ambulatory care and hospitalization after third-dose vaccination.** The four study arms included participants who previously received two-dose CoronaVac and were randomised to receive a third dose of CoronaVac ("CC-C") or BNT162b2 ("CC-B"), and participants who previously received two-dose BNT162b2 and were randomised to receive a third dose of CoronaVac ("BB-C") or BNT162b2 ("BB-B"). Reported discomfort associated with vaccination within the week after third-dose vaccination were highlighted in bold.

| | Study arm | Days between third-dose vaccination and seeking ambulatory care | Type of ambulatory care facility | Days between third-dose vaccination and hospitalization | Type of hospitalization facility | Summary |
|---|---|---|---|---|---|---|
| 1 | BB-B | 18 | Public Western medical practices | 18 | Public hospital | NOT related (haemorrhoids) |
| 2 | BB-C | 3 | Private Western medical practices | - | - | NOT related (nausea and dizziness) |
| 3 | BB-C | 5 | Public Western medical practices | - | - | NOT related (diarrhoea) |
| **4** | **CC-B** | **1** | **Other Chinese/ Western medical practices** | - | - | **Discomfort associated with vaccination within the week after vaccination (muscle pain)** |
| **5** | **CC-B** | **3** | **Private Western medical practices** | - | - | **Discomfort associated with vaccination within the week after vaccination (flu-like symptoms)** |
| **6** | **CC-B** | **5** | **Private Chinese medical practices** | - | - | **Discomfort associated with vaccination within the week after vaccination (flu-like symptoms & fatigue post-vaccination)** |
| 7 | CC-B | 15 | Public Western medical practices | - | - | NOT related (anaemia, normal ECG) |
| 8 | CC-B | 27 | Public Western medical practices | - | - | NOT related (flu) |
| **9** | **CC-C** | **7** | **Private Western medical practices** | - | - | **Discomfort associated with vaccination within the week after vaccination (muscle pain)** |
| 10 | CC-C | 24 | Public Western medical practices | - | - | NOT related (flu) |

**Table S5. Change in antibody and cell-mediated response after a randomised third dose of CoronaVac or BNT162b2 vaccine in adults who previously received two doses of either vaccine. (A) Change in antibody response on Day 28 compared to baseline.** The mean values for $PRNT_{50}$, $PRNT_{80}$ and $PRNT_{90}$ titers were considered as geometric mean titer (GMT), and the fold-change in PRNT titers on Day 28 from baseline were considered as geometric mean fold rise (GMFR). **Change in (B) IFNγ-producing $CD4^{+}$ and $CD8^{+}$ T cell responses against structural peptides of ancestral SARS-CoV-2 virus, and (C) their poly-cytokine production (TNFα and IL-2) and memory phenotype, on Day 7 and Day 28 compared to baseline.** The four study arms included participants who previously received two-dose CoronaVac and were randomised to receive a third dose of CoronaVac ("CC-C") or BNT162b2 ("CC-B"), and participants who previously received two-dose BNT162b2 and were randomised to receive a third dose of CoronaVac ("BB-C") or BNT162b2 ("BB-B"). Differences with p-values ≤ 0.05 were highlighted in bold. ELISA: enzyme-linked immunosorbent assay; sVNT: surrogate virus neutralisation test; PRNT: plaque reduction neutralisation test; ICS: intracellular cytokine staining; OD: optical density; % inhibition: percent inhibition; $PRNT_{50}$, $PRNT_{80}$ and $PRNT_{90}$: PRNT titers with endpoints at 50%, 80% and 90% inhibition respectively. Tn: naïve T cells; Tcm: central memory T cells; Tem: effector memory T cells; Teem: terminal effector memory T cells.

***Table S5A. Changes in antibody response on Day 28 compared to baseline.***

| | n | Baseline | Day 28 | Baseline vs. Day 28 | |
|---|---|---|---|---|---|
| | | **Mean** | **Mean** | **Fold change** | **p-value** |
| ***ELISA OD*** | | | | | |
| **Ancestral virus** | | | | | |
| CC-C | 100 | 0.38 | 1.73 | **4.5** | **<0.001** |
| CC-B | 115 | 0.39 | 2.43 | **6.3** | **<0.001** |
| BB-C | 111 | 1.31 | 1.8 | **1.4** | **<0.001** |
| BB-B | 109 | 1.35 | 2.35 | **1.7** | **<0.001** |
| ***sVNT % inhibition*** | | | | | |
| **Ancestral virus** | | | | | |
| CC-C | 100 | 17 | 83 | **5** | **<0.001** |
| CC-B | 115 | 16 | 96 | **6.1** | **<0.001** |
| BB-C | 111 | 66 | 87 | **1.3** | **<0.001** |
| BB-B | 109 | 70 | 96 | **1.4** | **<0.001** |
| **Omicron BA.1 variant** | | | | | |
| CC-C | 100 | 6 | 15 | **2.5** | **<0.001** |
| CC-B | 115 | 6 | 58 | **10.2** | **<0.001** |
| BB-C | 111 | 8 | 19 | 2.2 | **<0.001** |
| BB-B | 109 | 7 | 69 | **9.6** | **<0.001** |
| **Omicron BA.2 variant** | | | | | |
| CC-C | 100 | 5 | 43 | **8** | **<0.001** |
| CC-B | 115 | 6 | 85 | **13.6** | **<0.001** |
| BB-C | 111 | 28 | 50 | **1.8** | **<0.001** |
| BB-B | 109 | 29 | 90 | **3.1** | **<0.001** |
| ***$PRNT_{50}$ titer*** | | | | | |
| **Ancestral virus** | | | | | |
| CC-C | 20 | 8 | 109 | **13.5** | **<0.001** |
| CC-B | 20 | 10 | 905 | **93.7** | **<0.001** |
| BB-C | 20 | 34 | 92 | **2.7** | **<0.001** |
| BB-B | 20 | 43 | 816 | **19** | **<0.001** |
| **Omicron BA.1 variant** | | | | | |
| CC-C | 20 | 5 | 9 | **1.8** | **<0.01** |
| CC-B | 20 | 5 | 75 | **14.4** | **<0.001** |

| | | | | | | | | |
|---|---|---|---|---|---|---|---|---|
| BB-C | | 20 | | 6 | 8 | | **1.5** | **<0.01** |
| BB-B | | 20 | | 6 | 86 | | **15.5** | **<0.001** |
| **Omicron BA.2 variant** | | | | | | | | |
| CC-C | | 20 | | 6 | 6 | | 1.1 | 0.71 |
| CC-B | | 20 | | 5 | 80 | | **15.5** | **<0.001** |
| BB-C | | 20 | | 5 | 6 | | **1.2** | **0.02** |
| BB-B | | 20 | | 5 | 67 | | **12.6** | **<0.001** |
| ***$PRNT_{80}$ titer*** | | | | | | | | |
| **Ancestral virus** | | | | | | | | |
| CC-C | | 20 | | 5 | 57 | | **10.9** | **<0.001** |
| CC-B | | 20 | | 6 | 520 | | **84.4** | **<0.001** |
| BB-C | | 20 | | 17 | 46 | | **2.7** | **<0.001** |
| BB-B | | 20 | | 21 | 381 | | **17.8** | **<0.001** |
| **Omicron BA.1 variant** | | | | | | | | |
| CC-C | | 20 | | 5 | 6 | | **1.3** | **0.05** |
| CC-B | | 20 | | 5 | 39 | | **7.7** | **<0.001** |
| BB-C | | 20 | | 5 | 5 | | 1 | >0.99 |
| BB-B | | 20 | | 5 | 43 | | **8.6** | **<0.001** |
| **Omicron BA.2 variant** | | | | | | | | |
| CC-C | | 20 | | 5 | 6 | | 1 | >0.99 |
| CC-B | | 20 | | 5 | 41 | | **8.3** | **<0.001** |
| BB-C | | 20 | | 5 | 5 | | 1 | >0.99 |
| BB-B | | 20 | | 5 | 34 | | **6.5** | **<0.001** |
| ***$PRNT_{90}$ titer*** | | | | | | | | |
| **Ancestral virus** | | | | | | | | |
| CC-C | | 20 | | 5 | 31 | | **6.3** | **<0.001** |
| CC-B | | 20 | | 6 | 331 | | **59.7** | **<0.001** |
| BB-C | | 20 | | 10 | 25 | | **2.5** | **<0.001** |
| BB-B | | 20 | | 12 | 219 | | **18.4** | **<0.001** |
| **Omicron BA.1 variant** | | | | | | | | |
| CC-C | | 20 | | 5 | 6 | | 1.1 | 0.15 |
| CC-B | | 20 | | 5 | 22 | | **4.4** | **<0.001** |
| BB-C | | 20 | | 5 | 5 | | 1 | >0.99 |
| BB-B | | 20 | | 5 | 26 | | **5.3** | **<0.001** |
| **Omicron BA.2 variant** | | | | | | | | |
| CC-C | | 20 | | 5 | 5 | | 1 | 0.77 |
| CC-B | | 20 | | 5 | 25 | | **4.9** | **<0.001** |
| BB-C | | 20 | | 5 | 5 | | 1 | >0.99 |
| BB-B | | 20 | | 5 | 22 | | **4.4** | **<0.001** |

### *Table S5B. Changes in IFNγ-producing $CD4^+$ and $CD8^+$ T cell responses on Day 7 and Day 28 compared to baseline.*

| | **n** | | **Baseline** | **Day 7** | **Day 28** | | **Baseline vs. Day 7** | | | **Baseline vs. Day 28** | |
|---|---|---|---|---|---|---|---|---|---|---|---|
| | | | **Mean** | **Mean** | **Mean** | | **Fold change** | ***p*** | | **Fold change** | ***p*** |
| **IFNγ-producing $CD4^+$ cells (per 100,000)** | | | | | | | | | | | |
| CC-C | 20 | | 877 | 4248 | 2241 | | **4.8** | **<0.001** | | **2.6** | **0.04** |
| CC-B | 20 | | 641 | 3203 | 1822 | | **5** | **<0.01** | | **2.8** | **<0.01** |
| BB-C | 20 | | 3034 | 2745 | 2449 | | 0.9 | 0.9 | | 0.8 | 0.43 |
| BB-B | 20 | | 2196 | 2087 | 1383 | | 1 | 0.85 | | 0.6 | 0.6 |
| **IFNγ-producing $CD8^+$ cells (per 100,000)** | | | | | | | | | | | |
| CC-C | 20 | | 208 | 668 | 359 | | 3.2 | 0.13 | | 1.7 | 0.49 |
| CC-B | 20 | | 78 | 969 | 98 | | **12.4** | **<0.01** | | 1.2 | 0.53 |
| BB-C | 20 | | 213 | 122 | 218 | | 0.6 | 0.4 | | 1 | 0.94 |
| BB-B | 20 | | 146 | 173 | 164 | | 1.2 | 0.84 | | 1.1 | 0.97 |
| **IL4-producing $CD4^+$ cells (per 100,000)** | | | | | | | | | | | |
| CC-C | 20 | | 124 | 70 | 87 | | 0.6 | 0.38 | | 0.7 | 0.66 |
| CC-B | 20 | | 109 | 199 | 33 | | 1.8 | 0.28 | | **0.3** | **0.02** |

| | | | | | | | | | | | |
|---|---|---|---|---|---|---|---|---|---|---|---|
| BB-C | 20 | | 68 | 64 | 67 | | 0.9 | 0.95 | | 1 | 0.95 |
| BB-B | 20 | | 116 | 177 | 71 | | 1.5 | 0.37 | | 0.6 | 0.38 |

#### *Table S5C. Changes in poly-cytokine production (TNFα and IL-2) and memory phenotype in IFNγ-producing CD4$^+$ and CD8$^+$ T cells on Day 7 and Day 28 compared to baseline.*

| | n | Baseline | Day 7 | Day 28 | Baseline vs. Day 7 | | Baseline vs. Day 28 | |
|---|---|---|---|---|---|---|---|---|
| | | **Mean** | **Mean** | **Mean** | **Difference** | **p-value** | **Difference** | **p-value** |
| **Cytokine polyfunctional quality** | | | | | | | | |
| ***CD4$^+$ T cells*** | | | | | | | | |
| **CC-C** | 20 | | | | | | | |
| IFN$\gamma^+$ | | 45.1% | 58.8% | 53.3% | 13.7% | 0.23 | 8.2% | 0.51 |
| IFN$\gamma^+$ TNF$\alpha^+$ | | 11.5% | 23.4% | 8.1% | **11.9%** | **0.03** | -3.5% | 0.83 |
| IFN$\gamma^+$ IL2$^+$ | | 2.8% | 3.6% | 9.3% | 0.8% | 0.91 | **6.5%** | **0.02** |
| IFN$\gamma^+$ TNF$\alpha^+$ IL2$^+$ | | 10.6% | 14.2% | 9.3% | 3.6% | 0.13 | -1.3% | 0.7 |
| **CC-B** | 20 | | | | | | | |
| IFN$\gamma^+$ | | 42.6% | 61.7% | 61.6% | 19.1% | 0.14 | 19.0% | 0.07 |
| IFN$\gamma^+$ TNF$\alpha^+$ | | 8.1% | 21.4% | 9.0% | **13.3%** | **<0.01** | 1.0% | 0.32 |
| IFN$\gamma^+$ IL2$^+$ | | 2.2% | 3.1% | 3.6% | 0.9% | 0.73 | 1.4% | 0.25 |
| IFN$\gamma^+$ TNF$\alpha^+$ IL2$^+$ | | 7.2% | 8.8% | 5.9% | 1.6% | 0.43 | -1.3% | 0.66 |
| **BB-C** | 20 | | | | | | | |
| IFN$\gamma^+$ | | 51.2% | 50.2% | 47.2% | -1.0% | 0.72 | -4.0% | 0.48 |
| IFN$\gamma^+$ TNF$\alpha^+$ | | 15.0% | 11.1% | 10.6% | -3.9% | 0.18 | -4.4% | 0.18 |
| IFN$\gamma^+$ IL2$^+$ | | 5.4% | 10.7% | 11.6% | **5.4%** | **0.01** | **6.3%** | **0.02** |
| IFNg$^+$ TNF$\alpha^+$ IL2$^+$ | | 18.4% | 12.9% | 15.6% | -5.4% | 0.41 | -2.8% | 0.76 |
| **BB-B** | 20 | | | | | | | |
| IFN$\gamma^+$ | | 48.7% | 47.2% | 50.9% | -1.4% | 0.96 | 2.2% | 0.93 |
| IFN$\gamma^+$ TNF$\alpha^+$ | | 15.0% | 14.2% | 9.7% | -0.7% | 0.74 | -5.3% | 0.16 |
| IFN$\gamma^+$ IL2$^+$ | | 3.9% | 4.3% | 6.2% | 0.4% | 0.75 | 2.3% | 0.14 |
| IFN$\gamma^+$ TNF$\alpha^+$ IL2$^+$ | | 12.5% | 9.2% | 8.2% | -3.2% | 0.78 | -4.2% | 0.46 |
| ***CD8$^+$ T cells*** | | | | | | | | |
| **CC-C** | 20 | | | | | | | |
| IFN$\gamma^+$ | | 49.3% | 74.4% | 51.6% | 25.1% | 0.2 | 2.3% | 0.68 |
| IFN$\gamma^+$ TNF$\alpha^+$ | | 0.6% | 5.5% | 3.3% | **4.9%** | **0.02** | **2.7%** | **0.02** |
| IFN$\gamma^+$ IL2$^+$ | | 0.0% | 0.2% | 4.3% | 0.2% | >0.99 | **4.3%** | **0.04** |
| IFN$\gamma^+$ TNF$\alpha^+$ IL2$^+$ | | 0.1% | 0.0% | 0.8% | -0.1% | >0.99 | 0.7% | 0.42 |
| **CC-B** | 20 | | | | | | | |
| IFN$\gamma^+$ | | 27.5% | 71.0% | 43.2% | **43.5%** | **0.01** | 15.7% | 0.22 |
| IFN$\gamma^+$ TNF$\alpha^+$ | | 0.9% | 4.6% | 1.6% | 3.7% | 0.14 | 0.7% | 0.58 |
| IFN$\gamma^+$ IL2$^+$ | | 1.1% | 2.2% | 0.2% | 1.1% | 0.55 | -0.9% | 0.18 |
| IFN$\gamma^+$ TNF$\alpha^+$ IL2$^+$ | | 0.5% | 2.2% | 0.0% | 1.7% | 0.37 | -0.5% | >0.99 |
| **BB-C** | 20 | | | | | | | |
| IFN$\gamma^+$ | | 37.1% | 34.3% | 61.3% | -2.8% | 0.81 | 24.2% | 0.06 |

| | | | | | | | | | | | | |
|---|---|---|---|---|---|---|---|---|---|---|---|---|
| IFNγ$^{+}$ TNFα$^{+}$ | | | | 4.5% | 2.6% | 2.8% | | -2.0% | 0.8 | | -1.7% | 0.48 |
| IFNγ$^{+}$ IL2$^{+}$ | | | | 2.8% | 1.1% | 4.5% | | -1.7% | 0.58 | | 1.7% | 0.48 |
| IFNγ$^{+}$ TNFα$^{+}$ IL2$^{+}$ | | | | 5.6% | 2.1% | 1.3% | | -3.5% | 0.4 | | -4.3% | 0.31 |
| **BB-B** | | 20 | | | | | | | | | | |
| IFNγ$^{+}$ | | | | 30.3% | 39.8% | 41.2% | | 9.5% | 0.54 | | 11.0% | 0.58 |
| IFNγ$^{+}$ TNFα$^{+}$ | | | | 6.3% | 3.9% | 1.8% | | -2.4% | 0.77 | | -4.4% | 0.94 |
| IFNγ$^{+}$ IL2$^{+}$ | | | | 0.5% | 1.0% | 0.9% | | 0.6% | 0.58 | | 0.5% | 0.79 |
| IFNγ$^{+}$ TNFα$^{+}$ IL2$^{+}$ | | | | 3.0% | 0.2% | 1.0% | | -2.7% | 0.18 | | -2.0% | 0.79 |
| **Memory phenotype** | | | | | | | | | | | | |
| ***CD4$^{+}$ T cells*** | | | | | | | | | | | | |
| **CC-C** | | 20 | | | | | | | | | | |
| Tn | | | | 2.2% | 6.9% | 4.3% | | **4.7%** | **0.03** | | 2.1% | 0.14 |
| Teem | | | | 1.9% | 4.8% | 2.8% | | 2.9% | 0.1 | | 0.9% | 0.31 |
| Tcm | | | | 26.9% | 21.2% | 19.6% | | -5.7% | 0.48 | | -7.3% | 0.32 |
| Tem | | | | 39.1% | 67.1% | 53.3% | | **28.0%** | **<0.01** | | 14.2% | 0.11 |
| **CC-B** | | 20 | | | | | | | | | | |
| Tn | | | | 6.2% | 3.3% | 5.2% | | -2.9% | 0.13 | | -1.0% | 0.7 |
| Teem | | | | 1.4% | 0.9% | 3.2% | | -0.5% | 0.72 | | 1.8% | 0.26 |
| Tcm | | | | 15.3% | 26.2% | 20.6% | | **10.9%** | **<0.01** | | 5.3% | 0.12 |
| Tem | | | | 37.1% | 64.6% | 51.1% | | **27.5%** | **<0.01** | | 14.0% | 0.08 |
| **BB-C** | | 20 | | | | | | | | | | |
| Tn | | | | 7.2% | 2.4% | 4.5% | | -4.8% | 0.8 | | -2.6% | 0.49 |
| Teem | | | | 1.5% | 2.0% | 3.2% | | 0.5% | 0.6 | | 1.8% | 0.07 |
| Tcm | | | | 25.1% | 21.7% | 19.6% | | -3.4% | 0.41 | | -5.6% | 0.11 |
| Tem | | | | 56.2% | 58.9% | 57.6% | | 2.7% | 0.56 | | 1.4% | 0.56 |
| **BB-B** | | 20 | | | | | | | | | | |
| Tn | | | | 3.9% | 2.4% | 2.6% | | -1.5% | 0.62 | | -1.3% | >0.99 |
| Teem | | | | 1.1% | 1.4% | 2.0% | | 0.3% | >0.99 | | 0.9% | 0.53 |
| Tcm | | | | 24.7% | 16.8% | 18.8% | | **-7.9%** | **0.04** | | -5.9% | 0.36 |
| Tem | | | | 50.4% | 54.5% | 51.6% | | 4.1% | 0.24 | | 1.2% | 0.73 |
| ***CD8$^{+}$ T cells*** | | | | | | | | | | | | |
| **CC-C** | | 20 | | | | | | | | | | |
| Tn | | | | 11.5% | 14.1% | 10.3% | | 2.6% | 0.83 | | -1.2% | 0.64 |
| Teem | | | | 6.3% | 16.5% | 9.1% | | 10.2% | 0.11 | | 2.8% | 0.5 |
| Tcm | | | | 7.6% | 12.3% | 15.2% | | 4.7% | 0.29 | | 7.5% | 0.26 |
| Tem | | | | 24.6% | 37.1% | 25.4% | | 12.5% | 0.28 | | 0.8% | 0.8 |
| **CC-B** | | 20 | | | | | | | | | | |
| Tn | | | | 3.4% | 10.8% | 9.9% | | 7.4% | 0.04 | | 6.6% | 0.23 |
| Teem | | | | 4.5% | 9.2% | 4.6% | | 4.6% | 0.66 | | 0.1% | 0.83 |
| Tcm | | | | 5.9% | 19.1% | 11.0% | | **13.3%** | **0.01** | | 5.1% | 0.73 |
| Tem | | | | 16.2% | 40.9% | 19.5% | | **24.7%** | **0.02** | | 3.3% | >0.99 |

| | | | | | | | | | | | | |
|---|---|---|---|---|---|---|---|---|---|---|---|---|
| **BB-C** | | 20 | | | | | | | | | | |
| Tn | | | | 10.2% | 6.2% | 14.0% | | -4.1% | 0.33 | | 3.8% | 0.33 |
| Teem | | | | 7.2% | 3.1% | 10.9% | | -4.1% | 0.48 | | 3.7% | 0.72 |
| Tcm | | | | 10.7% | 6.6% | 12.2% | | -4.1% | 0.27 | | 1.5% | 0.75 |
| Tem | | | | 21.9% | 24.2% | 32.9% | | 2.3% | 0.94 | | 11.0% | 0.21 |
| **BB-B** | | 20 | | | | | | | | | | |
| Tn | | | | 9.1% | 4.5% | 8.1% | | -4.6% | 0.23 | | -1.1% | 0.62 |
| Teem | | | | 3.2% | 4.1% | 5.5% | | 0.9% | 0.67 | | 2.3% | 0.41 |
| Tcm | | | | 12.5% | 9.7% | 9.7% | | -2.9% | 0.96 | | -2.9% | 0.84 |
| Tem | | | | 15.2% | 26.8% | 21.8% | | 11.6% | 0.2 | | 6.7% | 0.37 |

**Table S6. Comparisons of antibody and cell-mediated response between a randomised third dose of CoronaVac or BNT162b2 vaccine in adults who previously received two doses of either vaccine. (A) Comparisons of antibody response on Day 28 between third dose CoronaVac or BNT162b2.** The mean values for $PRNT_{50}$, $PRNT_{80}$ and $PRNT_{90}$ titers were considered as geometric mean titer (GMT), and the fold-difference in PRNT titers between homologous and heterologous third dose were considered as geometric mean fold rise (GMFR). **Comparisons of (B) IFNγ-producing $CD4^{+}$ and $CD8^{+}$ T cell responses against structural peptides of ancestral SARS-CoV-2 virus, and (C) their poly-cytokine production (TNFα and IL-2) and memory phenotype, on Day 7 and 28 between third dose CoronaVac or BNT162b2.** The mean values of CD4+ or CD8+ T cells were expressed in number of cells per 100,000 cells. The four study arms included participants who previously received two-dose CoronaVac and were randomised to receive a third dose of CoronaVac ("CC-C") or BNT162b2 ("CC-B"), and participants who previously received two-dose BNT162b2 and were randomised to receive a third dose of CoronaVac ("BB-C") or BNT162b2 ("BB-B"). Differences with p-values ≤ 0.05 were highlighted in bold. ELISA: enzyme-linked immunosorbent assay; sVNT: surrogate virus neutralisation test; PRNT: plaque reduction neutralisation test; ICS: intracellular cytokine staining; OD: optical density; % inhibition: percent inhibition; $PRNT_{50}$, $PRNT_{80}$ and $PRNT_{90}$: PRNT titers with endpoints at 50%, 80% and 90% inhibition respectively; Tn: naïve T cells; Tcm: central memory T cells; Tem: effector memory T cells; Teem: terminal effector memory T cells.

***Table S6A. Comparisons of antibody response on Day 0 and 28 between third dose CoronaVac or BNT162b2.***

| | Baseline | | | | | Day 28 | | | |
|---|---|---|---|---|---|---|---|---|---|
| | **Third-dose CoronaVac** | **Third-dose BNT162b2** | **Fold-difference** | ***p*** | | **Third-dose CoronaVac** | **Third-dose BNT162b2** | **Fold-difference** | ***p*** |
| | **Mean** | **Mean** | | | | **Mean** | **Mean** | | |
| **<u>Prior two-dose CoronaVac</u>** | | | | | | | | | |
| **ELISA OD** | | | | | | | | | |
| Ancestral virus | 0.38 | 0.39 | 1 | 0.49 | | 1.73 | 2.43 | **1.4** | **<0.001** |
| **sVNT % inhibition** | | | | | | | | | |
| Ancestral virus | 17 | 16 | 0.9 | 0.62 | | 83 | 96 | **1.2** | **<0.001** |
| Omicron BA.1 variant | 6 | 6 | 0.9 | 0.38 | | 15 | 58 | **3.9** | **<0.001** |
| Omicron BA.2 variant | 5 | 6 | 1.2 | 0.15 | | 43 | 85 | **2** | **<0.001** |
| **PRNT50 titer** | | | | | | | | | |
| Ancestral virus | 8 | 10 | 1.2 | 0.55 | | 109 | 905 | **8.3** | **<0.001** |
| Omicron BA.1 variant | 5 | 5 | 1 | >0.99 | | 9 | 75 | **8** | **<0.001** |
| Omicron BA.2 variant | 6 | 5 | 0.9 | >0.99 | | 6 | 80 | **13** | **<0.001** |
| **PRNT80 titer** | | | | | | | | | |
| Ancestral virus | 5 | 6 | 1.2 | 0.15 | | 57 | 520 | **9.2** | **<0.001** |
| Omicron BA.1 variant | 5 | 5 | 1 | >0.99 | | 6 | 39 | **6.1** | **<0.001** |
| Omicron BA.2 variant | 5 | 5 | 0.9 | 0.34 | | 6 | 41 | **7.5** | **<0.001** |
| **PRNT90 titer** | | | | | | | | | |
| Ancestral virus | 5 | 6 | 1.1 | 0.08 | | 31 | 331 | **10.6** | **<0.001** |
| Omicron BA.1 variant | 5 | 5 | 1 | >0.99 | | 6 | 22 | **4** | **<0.001** |

| | | | | | | | | | |
|---|---|---|---|---|---|---|---|---|---|
| Omicron BA.2 variant | 5 | 5 | 1 | 0.34 | | 5 | 25 | **4.6** | **<0.001** |
| **Prior two-dose BNT162b2** | | | | | | | | | |
| **ELISA OD** | | | | | | | | | |
| Ancestral virus | 1.31 | 1.35 | 1 | 0.28 | | 1.8 | 2.35 | **1.3** | **<0.001** |
| **sVNT % inhibition** | | | | | | | | | |
| Ancestral virus | 66 | 70 | 1.1 | 0.17 | | 87 | 96 | **1.1** | **<0.001** |
| Omicron BA.1 variant | 8 | 7 | 0.9 | 0.23 | | 19 | 69 | **3.7** | **<0.001** |
| Omicron BA.2 variant | 28 | 29 | 1 | 0.42 | | 50 | 90 | **1.8** | **<0.001** |
| **PRNT50 titer** | | | | | | | | | |
| Ancestral virus | 34 | 43 | 1.3 | 0.21 | | 92 | 816 | **8.9** | **<0.001** |
| Omicron BA.1 variant | 6 | 6 | 1 | >0.99 | | 8 | 86 | **10.6** | **<0.001** |
| Omicron BA.2 variant | 5 | 5 | 1.1 | 0.34 | | 6 | 67 | **10.9** | **<0.001** |
| **PRNT80 titer** | | | | | | | | | |
| Ancestral virus | 17 | 21 | 1.3 | 0.21 | | 46 | 381 | **8.3** | **<0.001** |
| Omicron BA.1 variant | 5 | 5 | 1 | >0.99 | | 5 | 43 | **8.3** | **<0.001** |
| Omicron BA.2 variant | 5 | 5 | 1 | 0.34 | | 5 | 34 | **6.7** | **<0.001** |
| **PRNT90 titer** | | | | | | | | | |
| Ancestral virus | 10 | 12 | 1.2 | 0.31 | | 25 | 219 | **8.9** | **<0.001** |
| Omicron BA.1 variant | 5 | 5 | 1 | >0.99 | | 5 | 26 | **5.3** | **<0.001** |
| Omicron BA.2 variant | 5 | 5 | 1 | >0.99 | | 5 | 22 | **4.4** | **<0.001** |

#### *Table S6B. Comparisons of IFNγ-producing CD4⁺ and CD8⁺ T cell responses on Day 0, 7 and 28 between third dose CoronaVac or BNT162b2.*

| | **Baseline** | | | | **Day 7** | | | | **Day 28** | | | |
|---|---|---|---|---|---|---|---|---|---|---|---|---|
| | **Third-dose CoronaVac** | **Third-dose BNT162b2** | **Fold-difference** | ***p*** | **Third-dose CoronaVac** | **Third-dose BNT162b2** | **Fold-difference** | ***p*** | **Third-dose CoronaVac** | **Third-dose BNT162b2** | **Fold-difference** | ***p*** |
| | **Mean** | **Mean** | | | **Mean** | **Mean** | | | **Mean** | **Mean** | | |
| **Prior two-dose CoronaVac (per 100,000)** | | | | | | | | | | | | |
| IFNγ-producing CD4⁺ cells | 877 | 641 | 0.7 | 0.58 | 4248 | 3203 | 0.8 | 0.82 | 2241 | 1822 | 0.8 | 0.65 |
| IFNγ-producing CD8⁺ cells | 208 | 78 | 0.4 | 0.24 | 668 | 969 | 1.5 | 0.47 | 359 | 98 | 0.3 | 0.18 |
| IL4-producing CD4⁺ cells | 124 | 109 | 0.9 | 0.87 | 70 | 199 | 2.8 | 0.26 | 87 | 33 | 0.4 | 0.36 |
| **Prior two-dose BNT162b2 (per 100,000)** | | | | | | | | | | | | |
| IFNγ-producing CD4⁺ cells | 3034 | 2196 | 0.7 | 0.56 | 2745 | 2087 | 0.8 | 0.91 | 2449 | 1383 | 0.6 | 0.21 |
| IFNγ-producing CD8⁺ cells | 213 | 146 | 0.7 | 0.67 | 122 | 173 | 1.4 | 0.71 | 218 | 164 | 0.8 | 0.68 |
| IL4-producing CD4⁺ cells | 68 | 116 | 1.7 | 0.37 | 64 | 177 | 2.8 | 0.24 | 67 | 71 | 1.1 | 0.81 |

***Table S6C. Comparisons of poly-cytokine production (TNFα and IL-2) and memory phenotype in IFNγ-producing CD4⁺ and CD8⁺ T cells on Day 0, 7 and 28 between third dose CoronaVac or BNT162b2.***

| | **Baseline** | | | | **Day 7** | | | | **Day 28** | | | |
|---|---|---|---|---|---|---|---|---|---|---|---|---|
| | **Third-dose CoronaVac** | **Third-dose BNT162b2** | **Mean-difference** | **p-value** | **Third-dose CoronaVac** | **Third-dose BNT162b2** | **Mean-difference** | **p-value** | **Third-dose CoronaVac** | **Third-dose BNT162b2** | **Mean-difference** | **p-value** |
| | **Mean** | **Mean** | | | **Mean** | **Mean** | | | **Mean** | **Mean** | | |
| **Prior two-dose CoronaVac** | | | | | | | | | | | | |
| ***Cytokine polyfunctional quality*** | | | | | | | | | | | | |
| **CD4⁺ T cells** | | | | | | | | | | | | |
| IFNγ⁺ | 45.1% | 42.6% | -2.5% | 0.77 | 58.8% | 61.7% | 3.0% | 0.47 | 53.3% | 61.6% | 8.2% | 0.24 |
| IFNγ⁺ TNFα⁺ | 11.5% | 8.1% | -3.5% | 0.89 | 23.4% | 21.4% | -2.1% | 0.97 | 8.1% | 9.0% | 1.0% | 0.63 |
| IFNγ⁺ IL2⁺ | 2.8% | 2.2% | -0.6% | 0.92 | 3.6% | 3.1% | -0.5% | 0.97 | **9.3%** | **3.6%** | **-5.7%** | **0.01** |
| IFNγ⁺ TNFα⁺ IL2⁺ | 10.6% | 7.2% | -3.4% | 0.77 | 14.2% | 8.8% | -5.4% | 0.06 | 9.3% | 5.9% | -3.5% | 0.25 |
| **CD8⁺ T cells** | | | | | | | | | | | | |
| IFNγ⁺ | 49.3% | 27.5% | -21.8% | 0.09 | 74.4% | 71.0% | -3.4% | 0.52 | 51.6% | 43.2% | -8.4% | 0.94 |
| IFNγ⁺ TNFα⁺ | 0.6% | 0.9% | 0.3% | 0.65 | 5.5% | 4.6% | -0.9% | 0.57 | 3.3% | 1.6% | -1.7% | 0.11 |
| IFNγ⁺ IL2⁺ | 0.0% | 1.1% | 1.1% | 0.08 | 0.2% | 2.2% | 2.1% | 0.07 | **4.3%** | **0.2%** | **-4.0%** | **0.04** |
| IFNγ⁺ TNFα⁺ IL2⁺ | 0.1% | 0.5% | 0.4% | >0.99 | 0.0% | 2.2% | 2.2% | 0.16 | 0.8% | 0.0% | -0.8% | 0.16 |
| ***Memory phenotype*** | | | | | | | | | | | | |
| **CD4⁺ T cells** | | | | | | | | | | | | |
| Tn | **2.2%** | **6.2%** | **4.0%** | **0.05** | 6.9% | 3.3% | -3.6% | 0.25 | 4.3% | 5.2% | 0.8% | 0.3 |
| Teem | 1.9% | 1.4% | -0.5% | 0.54 | **4.8%** | **0.9%** | **-3.9%** | **0.01** | 2.8% | 3.2% | 0.4% | 0.79 |
| Tcm | 26.9% | 15.3% | -11.6% | 0.25 | 21.2% | 26.2% | 5.0% | 0.28 | 19.6% | 20.6% | 1.0% | 0.97 |
| Tem | 39.1% | 37.1% | -2.0% | 0.98 | 67.1% | 64.6% | -2.5% | 0.98 | 53.3% | 51.1% | -2.2% | 0.73 |
| **CD8⁺ T cells** | | | | | | | | | | | | |
| Tn | 11.5% | 3.4% | -8.1% | 0.13 | 14.1% | 10.8% | -3.3% | 0.66 | 10.3% | 9.9% | -0.4% | 0.31 |
| Teem | 6.3% | 4.5% | -1.7% | 0.92 | 16.5% | 9.2% | -7.3% | 0.08 | 9.1% | 4.6% | -4.5% | 0.28 |
| Tcm | 7.6% | 5.9% | -1.8% | >0.99 | 12.3% | 19.1% | 6.8% | 0.19 | 15.2% | 11.0% | -4.2% | 0.18 |
| Tem | 24.6% | 16.2% | -8.3% | 0.53 | 37.1% | 40.9% | 3.8% | 0.48 | 25.4% | 19.5% | -5.9% | 0.36 |
| **Prior two-dose BNT162b2** | | | | | | | | | | | | |
| ***Cytokine polyfunctional quality*** | | | | | | | | | | | | |
| **CD4⁺ T cells** | | | | | | | | | | | | |
| IFNγ⁺ | 51.2% | 48.7% | -2.6% | 0.8 | 50.2% | 47.2% | -3.0% | >0.99 | 47.2% | 50.9% | 3.6% | 0.37 |
| IFNγ⁺ TNFα⁺ | 15.0% | 15.0% | -0.1% | 0.9 | 11.1% | 14.2% | 3.1% | 0.27 | 10.6% | 9.7% | -0.9% | 0.43 |
| IFNγ⁺ IL2⁺ | 5.4% | 3.9% | -1.5% | 0.78 | **10.7%** | **4.3%** | **-6.4%** | **0.02** | **11.6%** | **6.2%** | **-5.4%** | **0.04** |
| IFNγ⁺ TNFα⁺ IL2⁺ | 18.4% | 12.5% | -5.9% | 0.15 | 12.9% | 9.2% | -3.7% | 0.3 | 15.6% | 8.2% | -7.3% | 0.08 |
| **CD8⁺ T cells** | | | | | | | | | | | | |

| | | | | | | | | | | | | |
|---|---|---|---|---|---|---|---|---|---|---|---|---|
| IFNγ+ | 37.1% | 30.3% | -6.8% | 0.5 | 34.3% | 39.8% | 5.5% | 0.66 | 61.3% | 41.2% | -20.1% | 0.34 |
| IFNγ+ TNFα+ | 4.5% | 6.3% | 1.7% | 0.89 | 2.6% | 3.9% | 1.3% | 0.73 | 2.8% | 1.8% | -1.0% | 0.52 |
| IFNγ+ IL2+ | 2.8% | 0.5% | -2.3% | 0.53 | 1.1% | 1.0% | 0.0% | >0.99 | 4.5% | 0.9% | -3.6% | 0.06 |
| IFNγ+ TNFα+ IL2+ | 5.6% | 3.0% | -2.6% | 0.43 | 2.1% | 0.2% | -1.8% | 0.53 | 1.3% | 1.0% | -0.3% | >0.99 |
| ***Memory phenotype*** | | | | | | | | | | | | |
| **CD4+ T cells** | | | | | | | | | | | | |
| Tn | 7.2% | 3.9% | -3.3% | 0.41 | 2.4% | 2.4% | 0.0% | 0.98 | 4.5% | 2.6% | -1.9% | 0.24 |
| Teem | 1.5% | 1.1% | -0.4% | 0.82 | 2.0% | 1.4% | -0.6% | 0.58 | 3.2% | 2.0% | -1.2% | 0.12 |
| Tcm | 25.1% | 24.7% | -0.5% | 0.94 | 21.7% | 16.8% | -5.0% | 0.36 | 19.6% | 18.8% | -0.7% | 0.8 |
| Tem | 56.2% | 50.4% | -5.8% | 0.68 | 58.9% | 54.5% | -4.4% | 0.94 | 57.6% | 51.6% | -6.1% | 0.61 |
| **CD8+ T cells** | | | | | | | | | | | | |
| Tn | 10.2% | 9.1% | -1.1% | 0.94 | 6.2% | 4.5% | -1.7% | >0.99 | 14.0% | 8.1% | -6.0% | 0.25 |
| Teem | 7.2% | 3.2% | -4.0% | 0.57 | 3.1% | 4.1% | 1.0% | 0.52 | 10.9% | 5.5% | -5.4% | 0.5 |
| Tcm | 10.7% | 12.5% | 1.9% | 0.86 | 6.6% | 9.7% | 3.1% | 0.6 | 12.2% | 9.7% | -2.5% | 0.55 |
| Tem | 21.9% | 15.2% | -6.7% | 0.54 | 24.2% | 26.8% | 2.6% | 0.81 | 32.9% | 21.8% | -11.1% | 0.21 |

**Table S7. Proportions of participants who were responders of cell-mediated response after a randomised third dose of CoronaVac or BNT162b2 vaccine in adults who previously received two doses of either vaccine, comparisons between a randomised third dose of CoronaVac or BNT162b2, and comparisons between Day 7 and Day 28 from baseline in each study arm.** The four study arms included participants who previously received two-dose CoronaVac and were randomised to receive a third dose of CoronaVac ("CC-C") or BNT162b2 ("CC-B"), and participants who previously received two-dose BNT162b2 and were randomised to receive a third dose of CoronaVac ("BB-C") or BNT162b2 ("BB-B"). Differences with p-values ≤ 0.05 were highlighted in bold.

| | **Baseline** | | | **Day 7** | | | **Day 28** | | | **Baseline vs. Day 7** | | **Baseline vs. Day 28** | |
|---|---|---|---|---|---|---|---|---|---|---|---|---|---|
| | **Third-dose CoronaVac** | **Third-dose BNT162b2** | ***p*** | **Third-dose CoronaVac** | **Third-dose BNT162b2** | ***p*** | **Third-dose CoronaVac** | **Third-dose BNT162b2** | ***p*** | **Third-dose CoronaVac** | **Third-dose BNT162b2** | **Third-dose CoronaVac** | **Third-dose BNT162b2** |
| | **n (%)** | **n (%)** | | **n (%)** | **n (%)** | | **n (%)** | **n (%)** | | ***p*** | ***p*** | ***p*** | ***p*** |
| **<u>Prior two-dose CoronaVac</u>** | | | | | | | | | | | | | |
| IFNγ-producing $CD4^+$ cells | 14 (70) | 12 (60) | 0.74 | 20 (100) | 19 (95) | >0.99 | 16 (80) | 16 (80) | >0.99 | **0.03** | **0.02** | 0.72 | 0.3 |
| IFNγ-producing $CD8^+$ cells | 10 (50) | 6 (30) | 0.33 | 16 (80) | 16 (80) | >0.99 | 12 (60) | 9 (45) | 0.53 | 0.1 | **<0.01** | 0.75 | 0.51 |
| IL4-producing $CD4^+$ cells | 13 (65) | 15 (75) | 0.73 | 12 (60) | 14 (70) | 0.74 | 13 (65) | 15 (75) | 0.73 | >0.99 | >0.99 | >0.99 | >0.99 |
| **<u>Prior two-dose BNT162b2</u>** | | | | | | | | | | | | | |
| IFNγ-producing $CD4^+$ cells | 18 (90) | 16 (80) | 0.66 | 17 (85) | 15 (75) | 0.69 | 17 (85) | 15 (75) | 0.69 | >0.99 | >0.99 | >0.99 | >0.99 |
| IFNγ-producing $CD8^+$ cells | 10 (50) | 8 (40) | 0.75 | 8 (40) | 9 (45) | >0.99 | 14 (70) | 9 (45) | 0.2 | 0.75 | >0.99 | 0.33 | >0.99 |
| IL4-producing $CD4^+$ cells | 12 (60) | 13 (65) | >0.99 | 12 (60) | 15 (75) | 0.5 | 12 (60) | 13 (65) | >0.99 | >0.99 | 0.73 | >0.99 | >0.99 |

**Table S8. List of trial participants with a SARS-CoV-2 infection identified by a RAT or PCR.** Altogether, (A) a total of 58 COVID-19 infection were identified either by RAT or PCR after third-dose vaccination until 31 May 2022 by various means, including (B) self-reported positive RAT or PCR result before the start of active surveillance, (C) during active surveillance, systematic monitoring for SARS-CoV-2 infection with self-administered RAT every 4 days, and (D) during active surveillance, upon a reported illness episode self-administered RAT daily starting from 4 days after the initiation of symptom diary. All COVID-19 RAT positive result identified during systematic monitoring and illness episodes were verified with the photo of the test kit whenever available. The four study arms included participants who previously received two-dose CoronaVac and were randomised to receive a third dose of CoronaVac ("CC-C") or BNT162b2 ("CC-B"), and participants who previously received two-dose BNT162b2 and were randomised to receive a third dose of CoronaVac ("BB-C") or BNT162b2 ("BB-B"). RAT: rapid antigen test; PCR: polymerase chain reaction. 1: Yes; 0: No; -: Unknown.

| | | | | A: Summary of SARS-CoV-2 infection identified | | B: Reported before initiation of active surveillance | | | C: Reported during active surveillance - systematic monitoring for SARS-CoV-2 infection | | | | | D: Reported during active surveillance - Daily symptom diary upon an illness episode | | | | |
|---|---|---|---|---|---|---|---|---|---|---|---|---|---|---|---|---|---|---|
| | Study arm | Date of third-dose vaccination (Month/ Year) | Date of COVID-19 infection (Month/ Year) | Positive verified with picture of RAT | Symptomatic illness | Positive by RAT or PCR | Positive by RAT | Positive by PCR | Positive by RAT or PCR | Positive by RAT | RAT result verified | Positive by PCR | Symptomatic illness | Positive by RAT or PCR | Positive by RAT | RAT result verified | Positive by PCR | Symptomatic illness |
| 1 | CC-C | 11/21 | 2/22 | 0 | 0 | 1 | 1 | 1 | 0 | 0 | - | - | - | - | - | - | - | - |
| 2 | CC-C | 11/21 | 3/22 | 1 | 1 | 1 | 1 | 1 | 1 | 1 | 1 | - | 1 | 1 | 0 | - | 1 | 1 |
| 3 | CC-B | 11/21 | 3/22 | 1 | 1 | 0 | 0 | 0 | 0 | 0 | 1 | 0 | 1 | 1 | 1 | 1 | - | 1 |
| 4 | CC-B | 11/21 | 3/22 | 1 | 1 | - | - | - | 0 | 0 | 1 | - | 1 | 1 | 1 | 1 | - | 1 |
| 5 | CC-B | 12/21 | 3/22 | 1 | 1 | 1 | 1 | - | 1 | 1 | 1 | - | 1 | 1 | 1 | 0 | 1 | 1 |
| 6 | CC-B | 12/21 | 3/22 | 1 | 1 | 1 | 1 | - | 1 | 1 | 0 | - | 1 | 1 | 1 | 1 | - | 1 |
| 7 | CC-B | 12/21 | 3/22 | 1 | 0 | 1 | 1 | 0 | 1 | 0 | 1 | 1 | 0 | 1 | 0 | - | 1 | 0 |
| 8 | BB-C | 12/21 | 3/22 | 1 | 0 | - | - | - | 1 | 1 | 1 | - | 0 | 0 | 0 | - | 0 | 0 |
| 9 | CC-B | 12/21 | 3/22 | 0 | 0 | 1 | 1 | 1 | 0 | 0 | - | 0 | - | - | - | - | - | - |
| 10 | CC-B | 12/21 | 2/22 | 0 | 0 | 1 | 1 | 1 | 0 | 0 | - | - | - | - | - | - | - | - |
| 11 | CC-B | 12/21 | 3/22 | 1 | 1 | - | 0 | - | 1 | 1 | 1 | 0 | 1 | 1 | 1 | 1 | 1 | 1 |
| 12 | CC-C | 12/21 | 2/22 | 0 | 0 | 1 | 1 | 1 | 0 | 0 | - | - | - | - | - | - | - | - |
| 13 | CC-C | 12/21 | 3/22 | 0 | 1 | - | 0 | - | 1 | 1 | 0 | - | 1 | 1 | 1 | 0 | - | 1 |
| 14 | BB-C | 12/21 | 2/22 | 0 | 0 | 1 | 1 | 1 | 0 | 0 | - | - | - | - | - | - | - | - |
| 15 | CC-C | 12/21 | 2/22 | 1 | 1 | 1 | 1 | 1 | 0 | 0 | 1 | 0 | 0 | 0 | 0 | - | 0 | 1 |
| 16 | CC-C | 12/21 | 2/22 | 0 | 0 | 1 | 1 | - | 0 | 0 | - | - | - | - | - | - | - | - |
| 17 | CC-C | 12/21 | 3/22 | 0 | 0 | 1 | 1 | 1 | 0 | 0 | - | - | - | - | - | - | - | - |
| 18 | CC-C | 12/21 | 2/22 | 0 | 0 | 1 | 1 | 1 | 0 | 0 | - | - | - | - | - | - | - | - |
| 19 | CC-C | 12/21 | 2/22 | 0 | 0 | 1 | 1 | 1 | 0 | 0 | - | - | - | - | - | - | - | - |
| 20 | CC-B | 12/21 | 2/22 | 0 | 0 | 1 | 1 | 0 | 0 | 0 | - | 0 | - | - | - | - | - | - |

| | | | | | | | | | | | | | | | | | | |
|---|---|---|---|---|---|---|---|---|---|---|---|---|---|---|---|---|---|---|
| 21 | CC-B | 12/21 | 3/22 | 1 | 1 | 1 | 1 | 1 | 0 | 0 | 1 | 0 | 1 | 1 | - | - | 1 | 1 |
| 22 | CC-B | 12/21 | 4/22 | 1 | 1 | - | 0 | - | 1 | 1 | 1 | - | 0 | 1 | 1 | 1 | - | 1 |
| 23 | CC-B | 12/21 | 3/22 | 1 | 1 | 1 | 1 | 1 | 1 | 1 | 1 | - | 1 | 1 | 1 | 1 | - | 1 |
| 24 | BB-C | 12/21 | 3/22 | 0 | 0 | 1 | 1 | - | 0 | 0 | - | - | - | - | - | - | - | - |
| 25 | BB-C | 12/21 | 3/22 | 1 | 1 | 0 | 0 | 0 | 1 | 1 | 1 | - | 0 | 1 | 1 | 1 | - | 1 |
| 26 | CC-C | 12/21 | 3/22 | 1 | 1 | 0 | 0 | 0 | 0 | 0 | 1 | 0 | 1 | 1 | - | 1 | - | 1 |
| 27 | CC-B | 12/21 | 3/22 | 1 | 1 | - | 0 | - | 1 | 1 | 1 | - | 1 | 0 | 0 | - | - | 1 |
| 28 | CC-C | 1/22 | 3/22 | 0 | 0 | 1 | 1 | - | 0 | - | - | - | - | - | - | - | - | - |
| 29 | BB-C | 1/22 | 4/22 | 1 | 1 | 0 | 0 | 0 | 1 | 1 | 1 | 1 | 1 | 1 | 1 | 1 | - | 1 |
| 30 | BB-B | 1/22 | 3/22 | 0 | 0 | 1 | 1 | - | 0 | 0 | - | 0 | - | - | - | - | - | - |
| 31 | BB-B | 1/22 | 3/22 | 0 | 0 | 1 | 1 | 0 | 0 | 0 | - | 0 | - | - | - | - | - | - |
| 32 | BB-B | 1/22 | 3/22 | 1 | 1 | 0 | 0 | 0 | 1 | 1 | 1 | 1 | 1 | 1 | 1 | 1 | - | 1 |
| 33 | BB-B | 1/22 | 3/22 | 1 | 1 | 0 | 0 | 0 | 0 | 0 | 1 | - | 1 | 1 | 1 | 1 | - | 1 |
| 34 | BB-B | 1/22 | 3/22 | 0 | 1 | 1 | 1 | 1 | 1 | 1 | 0 | 1 | 1 | 1 | 0 | - | 1 | 1 |
| 35 | BB-B | 1/22 | 3/22 | 0 | 1 | 1 | 1 | 1 | 1 | 1 | 0 | 1 | 1 | 1 | 0 | - | 1 | 1 |
| 36 | CC-B | 1/22 | 3/22 | 0 | 0 | 1 | 1 | 0 | 0 | 0 | - | - | - | - | - | - | - | - |
| 37 | BB-C | 1/22 | 2/22 | 0 | 0 | 1 | 0 | 1 | 0 | 0 | - | - | - | - | - | - | - | - |
| 38 | BB-C | 1/22 | 3/22 | 1 | 1 | 1 | 1 | 0 | 1 | 1 | 1 | - | 1 | 0 | 0 | - | - | 1 |
| 39 | CC-C | 1/22 | 3/22 | 0 | 0 | 1 | 1 | 0 | 0 | 0 | - | - | - | - | - | - | - | - |
| 40 | CC-C | 1/22 | 3/22 | 1 | 1 | 0 | 0 | 0 | 1 | 1 | 1 | - | 1 | 0 | 0 | - | - | 1 |
| 41 | BB-C | 1/22 | 2/22 | 0 | 0 | 1 | 0 | 1 | 0 | 0 | - | 0 | - | - | - | - | - | - |
| 42 | CC-B | 1/22 | 3/22 | 1 | 1 | 1 | - | 1 | 1 | 1 | 1 | - | 0 | 0 | 0 | - | 0 | 1 |
| 43 | BB-B | 1/22 | 3/22 | 1 | 1 | 1 | 1 | - | 1 | 1 | 1 | 1 | 1 | - | - | - | - | 1 |
| 44 | BB-B | 1/22 | 3/22 | 0 | 0 | 1 | 1 | 1 | 0 | 0 | - | 0 | - | - | - | - | - | - |
| 45 | BB-B | 1/22 | 3/22 | 1 | 1 | 1 | 1 | 0 | 1 | 1 | 1 | - | 1 | 0 | 0 | - | - | 1 |
| 46 | BB-C | 1/22 | 4/22 | 1 | 1 | 0 | 0 | 0 | 1 | 1 | 1 | 1 | 1 | 0 | 0 | - | 0 | 1 |
| 47 | BB-C | 1/22 | 2/22 | 0 | 0 | 1 | 1 | - | 0 | 0 | - | - | - | - | - | - | - | - |
| 48 | BB-C | 1/22 | 3/22 | 1 | 1 | 1 | 1 | - | 0 | 0 | 1 | 0 | 1 | 0 | 0 | - | 0 | 1 |
| 49 | BB-B | 1/22 | 3/22 | 1 | 1 | 1 | 1 | 0 | 1 | 1 | 1 | 0 | 1 | 1 | 0 | - | 1 | 1 |
| 50 | BB-B | 1/22 | 2/22 | 0 | 0 | 1 | 1 | - | 0 | 0 | - | - | - | - | - | - | - | - |
| 51 | BB-B | 1/22 | 5/22 | 1 | 1 | - | - | - | 1 | 1 | 1 | 1 | 1 | 1 | 0 | - | 1 | 1 |
| 52 | BB-C | 1/22 | 2/22 | 0 | 0 | 1 | 1 | 1 | 0 | 0 | - | - | - | - | - | - | - | - |
| 53 | BB-C | 1/22 | 2/22 | 0 | 0 | 1 | 1 | 1 | 0 | 0 | - | 0 | - | - | - | - | - | - |
| 54 | BB-C | 1/22 | 3/22 | 1 | 1 | 1 | 1 | 1 | 0 | 0 | 1 | - | 0 | - | - | - | - | 1 |
| 55 | BB-C | 1/22 | 3/22 | 1 | 1 | - | 0 | - | 0 | 0 | 1 | - | 0 | 1 | 0 | - | 1 | 1 |
| 56 | BB-C | 1/22 | 2/22 | 0 | 0 | 1 | 1 | 1 | 0 | 0 | - | 0 | - | - | - | - | - | - |
| 57 | CC-B | 1/22 | 3/22 | 0 | 0 | 1 | 1 | 1 | 0 | 0 | - | 0 | - | - | - | - | - | - |
| 58 | BB-B | 1/22 | 3/22 | 1 | 1 | 1 | 1 | 0 | 1 | 1 | 1 | - | 1 | 1 | 0 | - | 1 | 1 |

**Table S9. (A) Participant characteristics, as well as (B) antibody and cell-mediated responses of all vaccinated participants at baseline, stratified by prior two-dose CoronaVac or BNT162b2.** We enrolled 219 adults who previously received two doses of CoronaVac and 232 adults who previously received two doses of BNT162b2, and randomly assigned participants to receive either vaccine as a third dose in each group. Differences with p-values ≤ 0.05 were highlighted in bold. ELISA: enzyme-linked immunosorbent assay; sVNT: surrogate virus neutralisation test; PRNT: plaque reduction neutralisation test; ICS: intracellular cytokine staining; OD: optical density; % inhibition: percent inhibition; $PRNT_{50}$, $PRNT_{80}$ and $PRNT_{90}$: PRNT titers with endpoints at 50%, 80% and 90% inhibition respectively.

***Table S9A. Participant characteristics at baseline.***

| | | **Received primary series of CoronaVac (CC)** | **Received primary series of BNT162b2 (BB)** | | ***p*** |
|---|---|---|---|---|---|
| | | **(n = 219)** | **(n = 232)** | | |
| | | **n (%)** | **n (%)** | | |
| **Female** | | 111 (51) | 110 (47) | | >0.99 |
| **Age (median, IQR)** | | 55 (47-61) | 54 (46-60) | | 0.99 |
| **Age group (years)** | | | | | |
| <18 | | 0 (0) | 0 (0) | | |
| 18 - 29 | | 10 (5) | 12 (5) | | |
| 30 - 39 | | 21 (10) | 17 (7) | | |
| 40 - 49 | | 47 (21) | 53 (23) | | |
| 50 - 59 | | 81 (37) | 92 (40) | | |
| 60 - 69 | | 53 (24) | 46 (20) | | |
| 70 - 79 | | 7 (3) | 12 (5) | | |
| ≥80 | | 0 (0) | 0 (0) | | |
| **Chinese** | | 216 (99) | 227 (98) | | 0.63 |
| **Obesity** | | | | | |
| Underweight | | 12 (5) | 11 (5) | | 0.49 |
| Normal | | 97 (44) | 90 (39) | | |
| Overweight | | 42 (19) | 57 (25) | | |
| Obese | | 68 (31) | 74 (32) | | |
| **Chronic medical conditions** | | | | | |
| Any | | 42 (19) | 62 (27) | | 0.06 |
| Lung disease, including COPD and asthma | | 2 (1) | 1 (0) | | >0.99 |
| Heart disease | | 3 (1) | 2 (1) | | >0.99 |
| Hypertension | | 19 (9) | 26 (11) | | 0.37 |
| Diabetes | | 8 (4) | 5 (2) | | 0.58 |
| Hypercholesterolemia | | 10 (5) | 18 (8) | | 0.18 |
| Kidney disease | | 0 (0) | 1 (0) | | >0.99 |
| Liver disease | | 3 (1) | 4 (2) | | >0.99 |
| Cancer | | 5 (2) | 1 (0) | | 0.22 |
| **Days between first and second dose of COVID-19 vaccination (median, IQR)** | | **28 (28-29)** | **21 (21-23)** | | **<0.01** |
| **Days between second and third (study) dose of COVID-19 vaccination (median, IQR)** | | **236 (213-247)** | **224 (195-246)** | | **0.02** |
| **Smoking** | | | | | |
| Ever | | **15 (7)** | **5 (2)** | | **0.04** |
| Current | | **9 (4)** | **2 (1)** | | **0.07** |

***Table S9B. Antibody and cell-mediated response at baseline.***

| | Received primary series of CoronaVac (CC) | Received primary series of BNT162b2 (BB) | Fold-difference | *p* |
|---|---|---|---|---|
| | **(n = 219)** | **(n = 232)** | | |
| | **Mean** | **Mean** | | |
| **Antibody response** | | | | |
| ***ELISA OD*** | | | | |
| Ancestral virus | **0.38** | **1.33** | **3.5** | **<0.01** |
| ***sVNT % inhibition*** | | | | |
| Ancestral virus | **16** | **68** | **4.2** | **<0.01** |
| Omicron BA.1 variant | **6** | **8** | **1.3** | **<0.01** |
| Omicron BA.2 variant | **6** | **29** | **4.9** | **<0.01** |
| ***PRNT50 titer*** | | | | |
| Ancestral virus | **9** | **38** | **4.3** | **<0.01** |
| Omicron BA.1 variant | 5 | 6 | 1.1 | 0.14 |
| Omicron BA.2 variant | 5 | 5 | 1.0 | 0.57 |
| ***PRNT80 titer*** | | | | |
| Ancestral virus | **6** | **19** | **3.4** | **<0.01** |
| Omicron BA.1 variant | 5 | 5 | 1.0 | |
| Omicron BA.2 variant | 5 | 5 | 1.0 | >0.99 |
| ***PRNT90 titer*** | | | | |
| Ancestral virus | **5** | **11** | **2.1** | **<0.01** |
| Omicron BA.1 variant | 5 | 5 | 1.0 | |
| Omicron BA.2 variant | 5 | 5 | 1.0 | 0.33 |
| **Cell-mediated response** | | | | |
| IFNγ-producing $CD4^+$ responder (n, %) | 26 (65) | 34 (85) | - | 0.07 |
| IFNγ-producing $CD8^+$ responder (n, %) | 16 (40) | 18 (45) | - | 0.82 |
| IL4-producing $CD4^+$ cells responder (n, %) | 28 (70) | 25 (63) | - | 0.64 |
| IFNγ-producing $CD4^+$ cells (per 100,000) | **750** | **2581** | **3.4** | **<0.01** |
| IFNγ-producing $CD8^+$ cells (per 100,000) | 128 | 176 | 1.4 | 0.58 |
| IL4-producing $CD4^+$ cells (per 100,000) | 116 | 89 | 0.8 | 0.73 |

### Supplementary Figures

**Figure S1. A detailed flow chart of participant enrolment for this open-label randomised trial of CoronaVac or BNT162b2 provided as a third vaccine dose in adults who previously received two doses of either vaccine, including reasons for exclusion at each stage. (A)** From eligibility assessment for enrolment to randomisation of study intervention. **(B)** From randomisation to administration of study intervention. **(C)** From administration of study intervention to selection of paired Day 0 and Day 28 blood samples for laboratory testing.

**A**

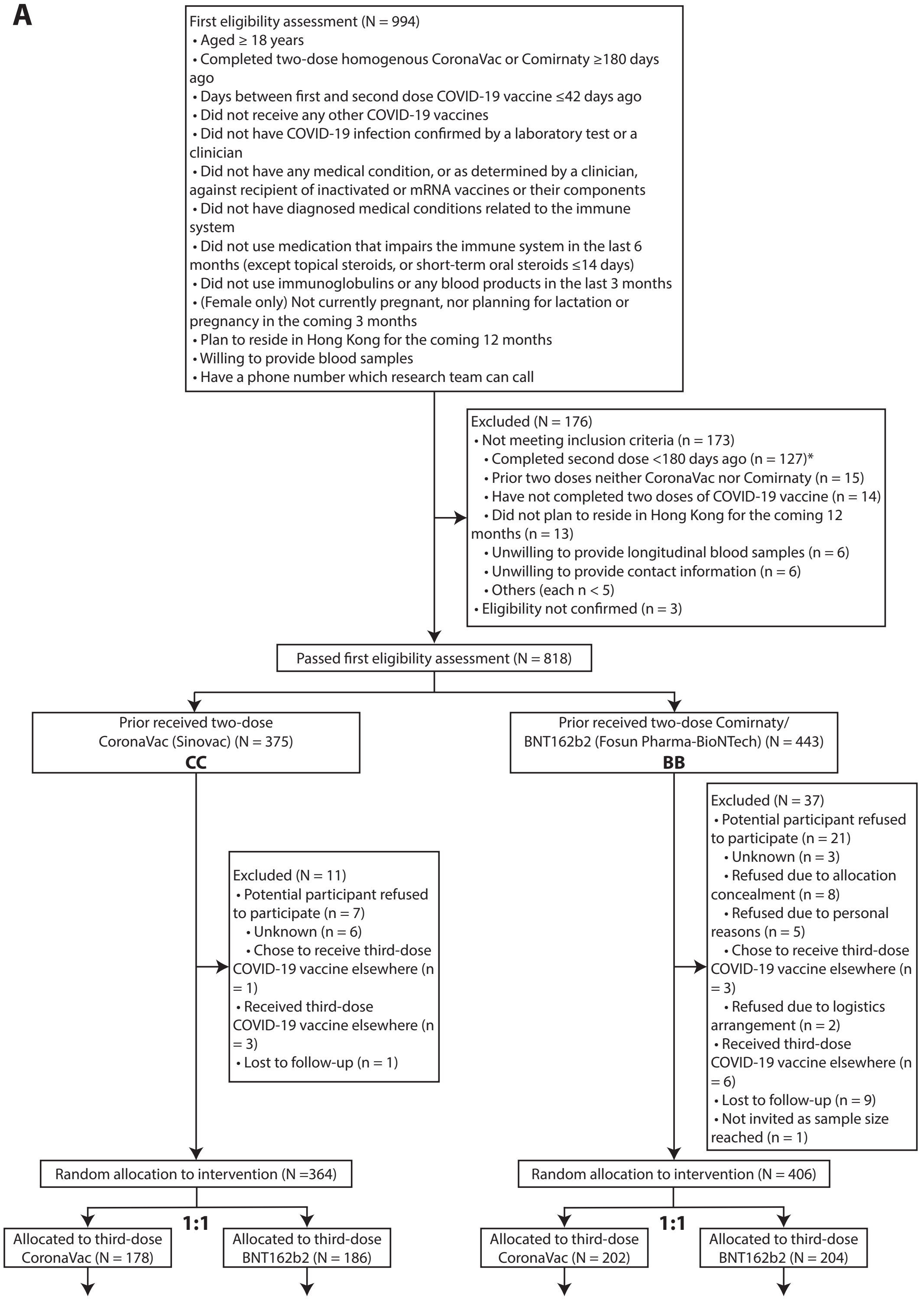

B

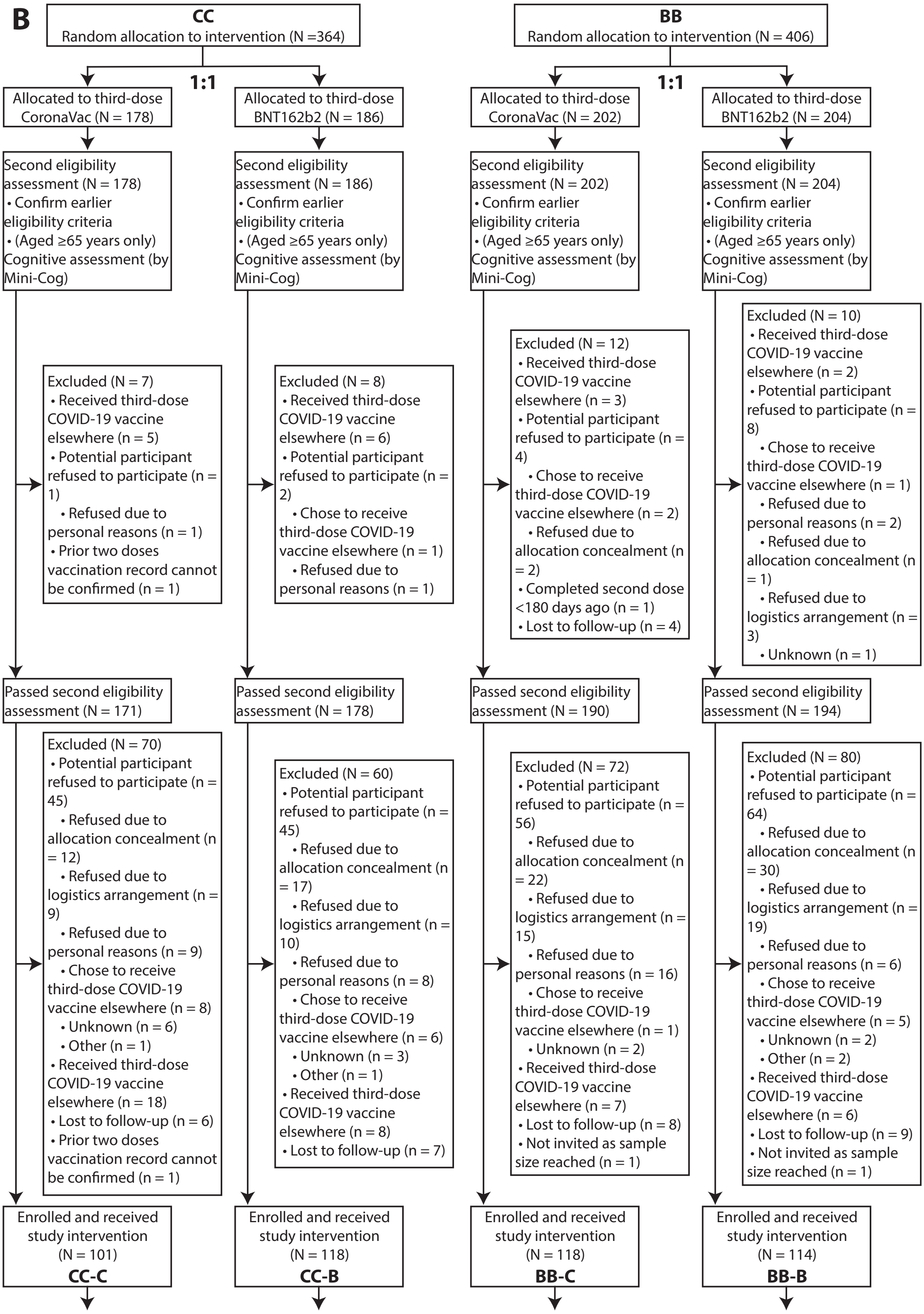
CC
Random allocation to intervention (N =364)
1:1
Allocated to third-dose CoronaVac (N = 178)
Allocated to third-dose BNT162b2 (N = 186)
Second eligibility assessment (N = 178)
• Confirm earlier eligibility criteria
• (Aged ≥65 years only) Cognitive assessment (by Mini-Cog)
Second eligibility assessment (N = 186)
• Confirm earlier eligibility criteria
• (Aged ≥65 years only) Cognitive assessment (by Mini-Cog)
Excluded (N = 7)
• Received third-dose COVID-19 vaccine elsewhere (n = 5)
• Potential participant refused to participate (n = 1)
• Refused due to personal reasons (n = 1)
• Prior two doses vaccination record cannot be confirmed (n = 1)
Excluded (N = 8)
• Received third-dose COVID-19 vaccine elsewhere (n = 6)
• Potential participant refused to participate (n = 2)
• Chose to receive third-dose COVID-19 vaccine elsewhere (n = 1)
• Refused due to personal reasons (n = 1)
Passed second eligibility assessment (N = 171)
Passed second eligibility assessment (N = 178)
Excluded (N = 70)
• Potential participant refused to participate (n = 45)
• Refused due to allocation concealment (n = 12)
• Refused due to logistics arrangement (n = 9)
• Refused due to personal reasons (n = 9)
• Chose to receive third-dose COVID-19 vaccine elsewhere (n = 8)
• Unknown (n = 6)
• Other (n = 1)
• Received third-dose COVID-19 vaccine elsewhere (n = 18)
• Lost to follow-up (n = 6)
• Prior two doses vaccination record cannot be confirmed (n = 1)
Excluded (N = 60)
• Potential participant refused to participate (n = 45)
• Refused due to allocation concealment (n = 17)
• Refused due to logistics arrangement (n = 10)
• Refused due to personal reasons (n = 8)
• Chose to receive third-dose COVID-19 vaccine elsewhere (n = 6)
• Unknown (n = 3)
• Other (n = 1)
• Received third-dose COVID-19 vaccine elsewhere (n = 8)
• Lost to follow-up (n = 7)
Enrolled and received study intervention (N = 101)
CC-C
Enrolled and received study intervention (N = 118)
CC-B
BB
Random allocation to intervention (N = 406)
1:1
Allocated to third-dose CoronaVac (N = 202)
Allocated to third-dose BNT162b2 (N = 204)
Second eligibility assessment (N = 202)
• Confirm earlier eligibility criteria
• (Aged ≥65 years only) Cognitive assessment (by Mini-Cog)
Second eligibility assessment (N = 204)
• Confirm earlier eligibility criteria
• (Aged ≥65 years only) Cognitive assessment (by Mini-Cog)
Excluded (N = 12)
• Received third-dose COVID-19 vaccine elsewhere (n = 3)
• Potential participant refused to participate (n = 4)
• Chose to receive third-dose COVID-19 vaccine elsewhere (n = 2)
• Refused due to allocation concealment (n = 2)
• Completed second dose <180 days ago (n = 1)
• Lost to follow-up (n = 4)
Excluded (N = 10)
• Received third-dose COVID-19 vaccine elsewhere (n = 2)
• Potential participant refused to participate (n = 8)
• Chose to receive third-dose COVID-19 vaccine elsewhere (n = 1)
• Refused due to personal reasons (n = 2)
• Refused due to allocation concealment (n = 1)
• Refused due to logistics arrangement (n = 3)
• Unknown (n = 1)
Passed second eligibility assessment (N = 190)
Passed second eligibility assessment (N = 194)
Excluded (N = 72)
• Potential participant refused to participate (n = 56)
• Refused due to allocation concealment (n = 22)
• Refused due to logistics arrangement (n = 15)
• Refused due to personal reasons (n = 16)
• Chose to receive third-dose COVID-19 vaccine elsewhere (n = 1)
• Unknown (n = 2)
• Received third-dose COVID-19 vaccine elsewhere (n = 7)
• Lost to follow-up (n = 8)
• Not invited as sample size reached (n = 1)
Excluded (N = 80)
• Potential participant refused to participate (n = 64)
• Refused due to allocation concealment (n = 30)
• Refused due to logistics arrangement (n = 19)
• Refused due to personal reasons (n = 6)
• Chose to receive third-dose COVID-19 vaccine elsewhere (n = 5)
• Unknown (n = 2)
• Other (n = 2)
• Received third-dose COVID-19 vaccine elsewhere (n = 6)
• Lost to follow-up (n = 9)
• Not invited as sample size reached (n = 1)
Enrolled and received study intervention (N = 118)
BB-C
Enrolled and received study intervention (N = 114)
BB-B

C

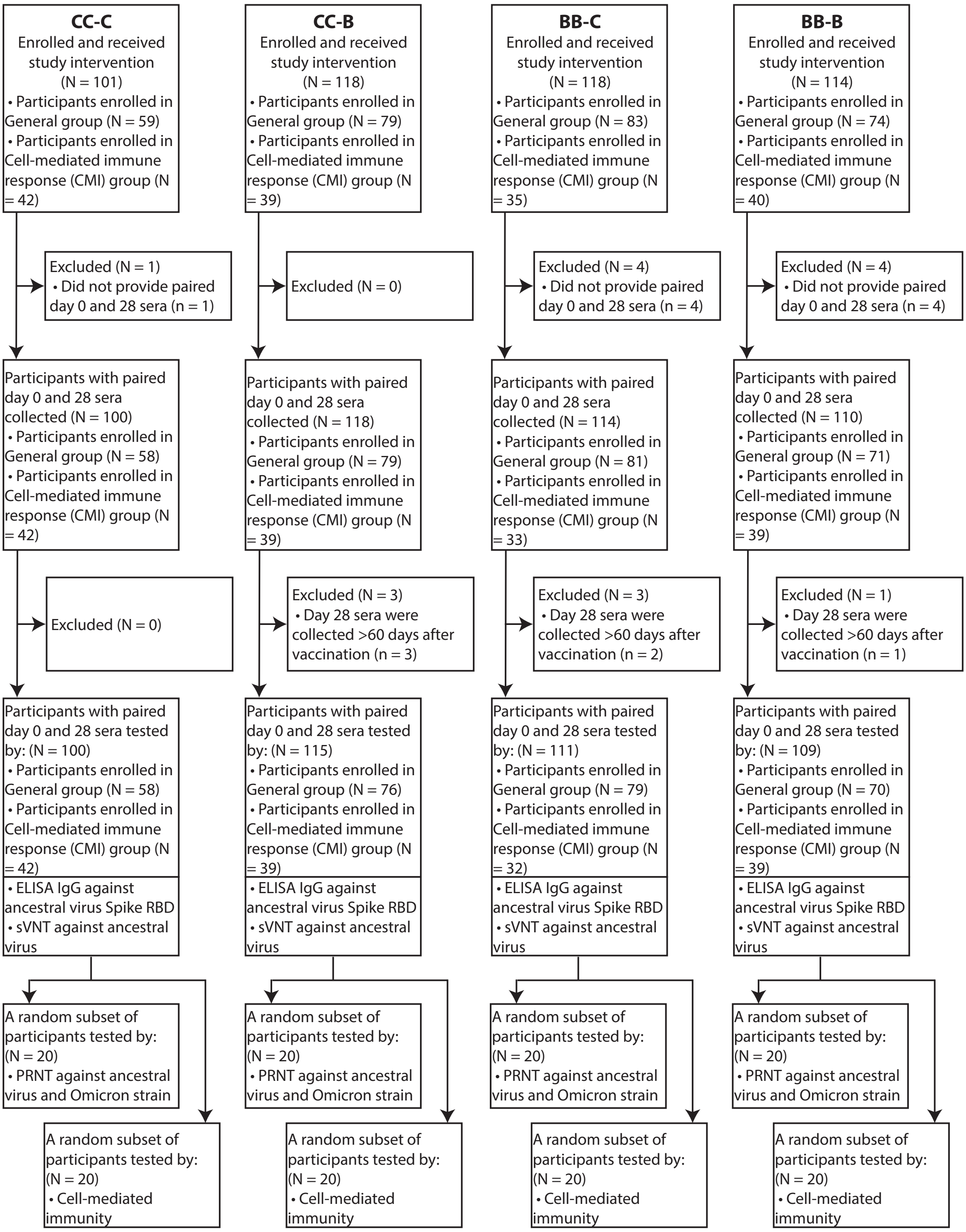
CC-C
Enrolled and received study intervention (N = 101)
• Participants enrolled in General group (N = 59)
• Participants enrolled in Cell-mediated immune response (CMI) group (N = 42)
Excluded (N = 1)
• Did not provide paired day 0 and 28 sera (n = 1)
Participants with paired day 0 and 28 sera collected (N = 100)
• Participants enrolled in General group (N = 58)
• Participants enrolled in Cell-mediated immune response (CMI) group (N = 42)
Excluded (N = 0)
Participants with paired day 0 and 28 sera tested by: (N = 100)
• Participants enrolled in General group (N = 58)
• Participants enrolled in Cell-mediated immune response (CMI) group (N = 42)
• ELISA IgG against ancestral virus Spike RBD
• sVNT against ancestral virus
A random subset of participants tested by: (N = 20)
• PRNT against ancestral virus and Omicron strain
A random subset of participants tested by: (N = 20)
• Cell-mediated immunity
CC-B
Enrolled and received study intervention (N = 118)
• Participants enrolled in General group (N = 79)
• Participants enrolled in Cell-mediated immune response (CMI) group (N = 39)
Excluded (N = 0)
Participants with paired day 0 and 28 sera collected (N = 118)
• Participants enrolled in General group (N = 79)
• Participants enrolled in Cell-mediated immune response (CMI) group (N = 39)
Excluded (N = 3)
• Day 28 sera were collected >60 days after vaccination (n = 3)
Participants with paired day 0 and 28 sera tested by: (N = 115)
• Participants enrolled in General group (N = 76)
• Participants enrolled in Cell-mediated immune response (CMI) group (N = 39)
• ELISA IgG against ancestral virus Spike RBD
• sVNT against ancestral virus
A random subset of participants tested by: (N = 20)
• PRNT against ancestral virus and Omicron strain
A random subset of participants tested by: (N = 20)
• Cell-mediated immunity
BB-C
Enrolled and received study intervention (N = 118)
• Participants enrolled in General group (N = 83)
• Participants enrolled in Cell-mediated immune response (CMI) group (N = 35)
Excluded (N = 4)
• Did not provide paired day 0 and 28 sera (n = 4)
Participants with paired day 0 and 28 sera collected (N = 114)
• Participants enrolled in General group (N = 81)
• Participants enrolled in Cell-mediated immune response (CMI) group (N = 33)
Excluded (N = 3)
• Day 28 sera were collected >60 days after vaccination (n = 2)
Participants with paired day 0 and 28 sera tested by: (N = 111)
• Participants enrolled in General group (N = 79)
• Participants enrolled in Cell-mediated immune response (CMI) group (N = 32)
• ELISA IgG against ancestral virus Spike RBD
• sVNT against ancestral virus
A random subset of participants tested by: (N = 20)
• PRNT against ancestral virus and Omicron strain
A random subset of participants tested by: (N = 20)
• Cell-mediated immunity
BB-B
Enrolled and received study intervention (N = 114)
• Participants enrolled in General group (N = 74)
• Participants enrolled in Cell-mediated immune response (CMI) group (N = 40)
Excluded (N = 4)
• Did not provide paired day 0 and 28 sera (n = 4)
Participants with paired day 0 and 28 sera collected (N = 110)
• Participants enrolled in General group (N = 71)
• Participants enrolled in Cell-mediated immune response (CMI) group (N = 39)
Excluded (N = 1)
• Day 28 sera were collected >60 days after vaccination (n = 1)
Participants with paired day 0 and 28 sera tested by: (N = 109)
• Participants enrolled in General group (N = 70)
• Participants enrolled in Cell-mediated immune response (CMI) group (N = 39)
• ELISA IgG against ancestral virus Spike RBD
• sVNT against ancestral virus
A random subset of participants tested by: (N = 20)
• PRNT against ancestral virus and Omicron strain
A random subset of participants tested by: (N = 20)
• Cell-mediated immunity

**Figure S2. Boosting of (A) neutralising antibody response and (B) cell-mediated response against ancestral SARS-CoV-2 virus, Omicron BA.1 and BA.2 variants at Day 7 and/or Day 28 after a randomised third dose of CoronaVac or BNT162b2 vaccine in adults who previously received two doses of either vaccine, measured by live virus plaque reduction neutralization test (PRNT) or intracellular cytokine staining (ICS) respectively.** PRNT titers were evaluated with endpoint at 50% inhibition (PRNT50). The four study arms included participants who previously received two-dose CoronaVac and were randomised to receive a third dose of CoronaVac ("CC-C") or BNT162b2 ("CC-B"), and participants who previously received two-dose BNT162b2 and were randomised to receive a third dose of CoronaVac ("BB-C") or BNT162b2 ("BB-B"). The symbol X and the numbers above each panel indicate the fold-change compared to baseline (Day 0). D28/D0: comparison between Day 28 against Day 0; D7/D0: comparison between Day 7 against Day 0.

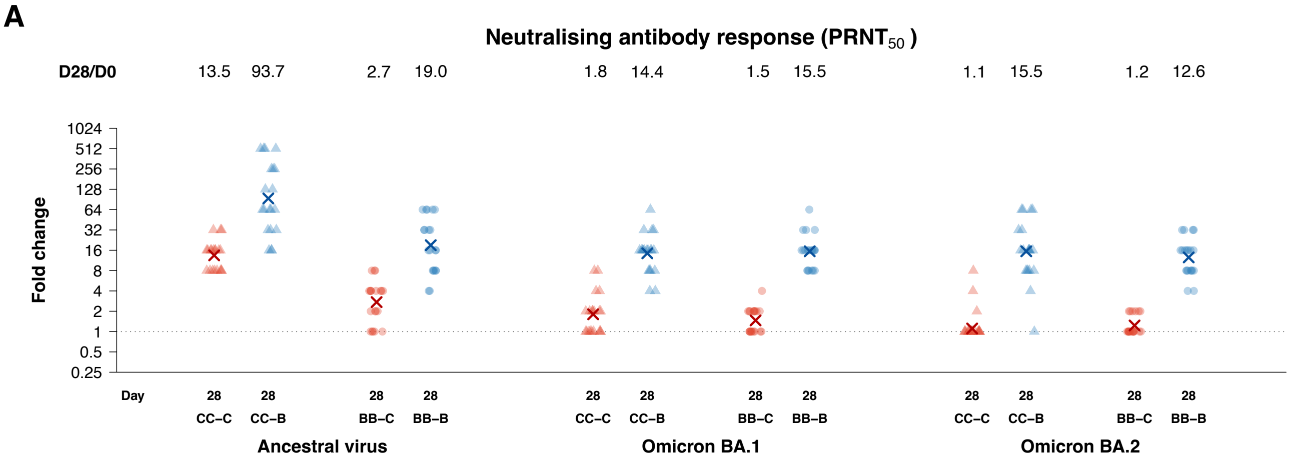

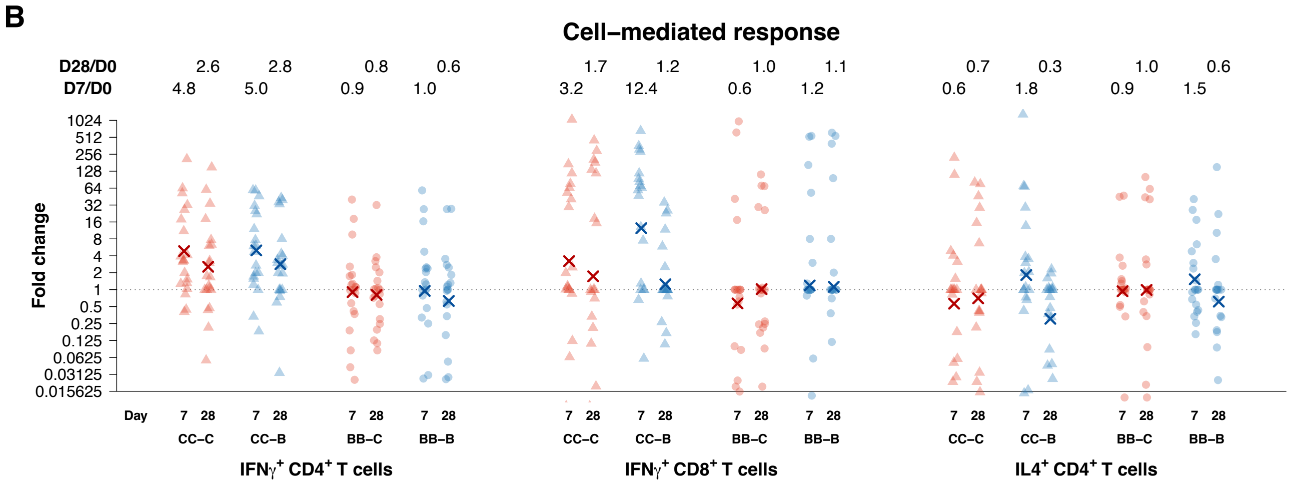

**Figure S3. Serum antibodies against ancestral SARS-CoV-2 virus at baseline and 28 days after a randomised third dose of CoronaVac or BNT162b2 vaccine in adults who previously received two doses of either vaccine, measured by (A) ELISA, (B) surrogate virus neutralization test (sVNT) or (C) live virus plaque reduction neutralization test (PRNT).** PRNT titers were evaluated with endpoint at 50% inhibition (PRNT50). The four study arms included participants who previously received two-dose CoronaVac and were randomised to receive a third dose of CoronaVac ("CC-C") or BNT162b2 ("CC-B"), and participants who previously received two-dose BNT162b2 and were randomised to receive a third dose of CoronaVac ("BB-C") or BNT162b2 ("BB-B"). The symbol X and the numbers above each panel indicate the mean level. p-values ≤ 0.05 *, ≤ 0.01 **, ≤ 0.001 ***

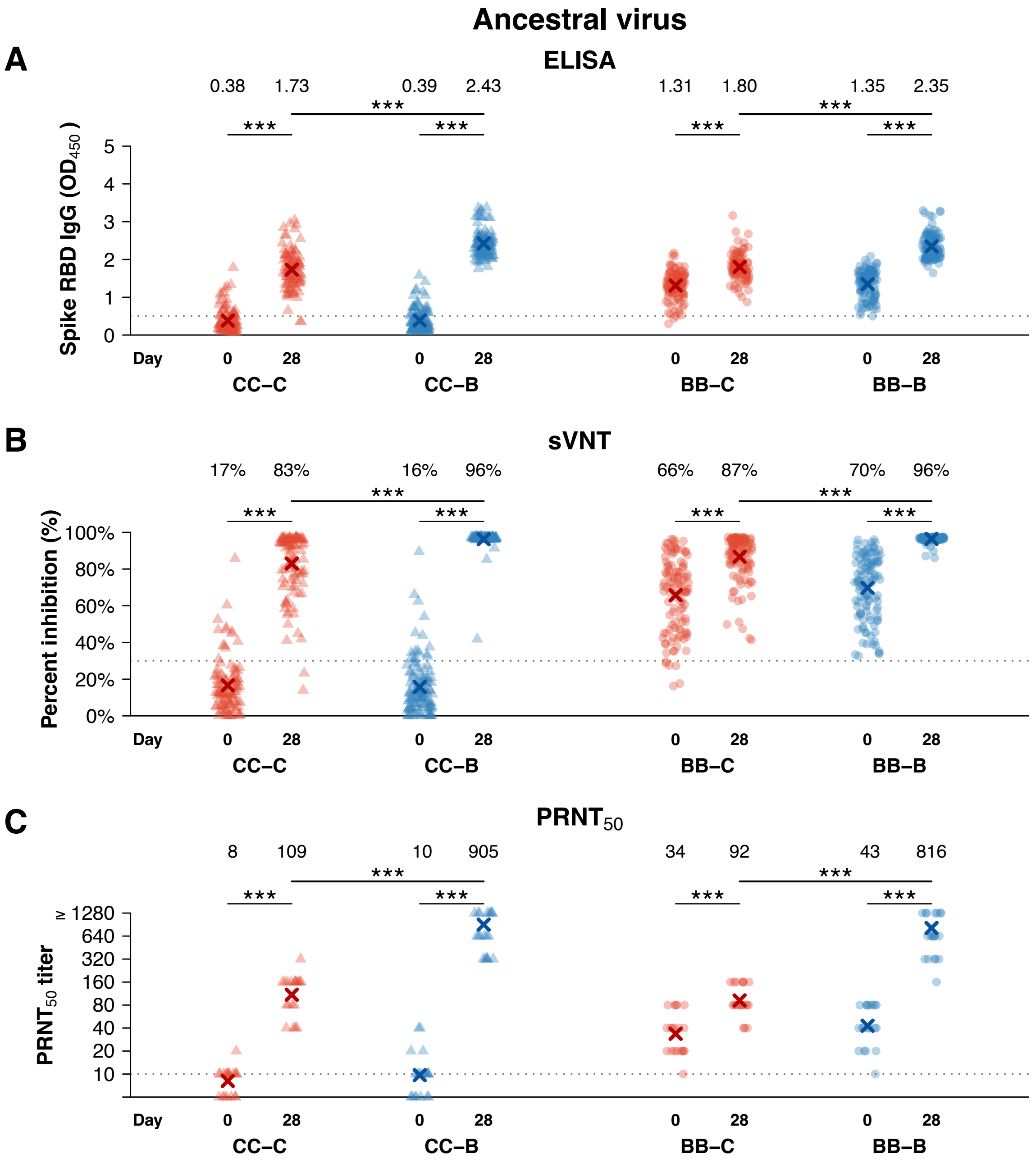

**Figure S4. Serum neutralising antibodies against (A-C) ancestral SARS-CoV-2 virus, (D-F) Omicron BA.1 and (G-I) Omicron BA.2 variants at baseline and 28 days after a randomised third dose of CoronaVac or BNT162b2 vaccine in adults who previously received two doses of either vaccine, measured by live virus plaque reduction neutralization test (PRNT) with endpoints at 50% (PRNT50), 80% (PRNT80) and 90% (PRNT90) respectively.** The four study arms included participants who previously received two-dose CoronaVac and were randomised to receive a third dose of CoronaVac ("CC-C") or BNT162b2 ("CC-B"), and participants who previously received two-dose BNT162b2 and were randomised to receive a third dose of CoronaVac ("BB-C") or BNT162b2 ("BB-B"). The symbol X and the numbers above each panel indicate the mean level.

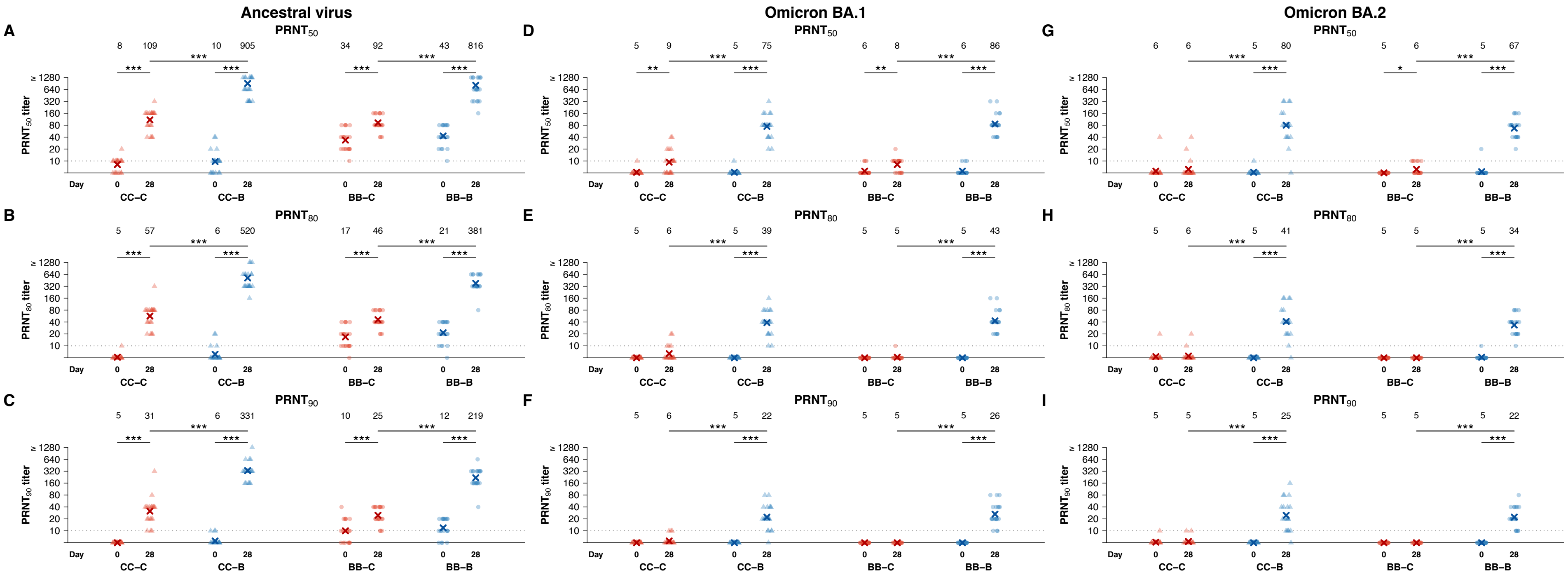

**Figure S5. IL-4 producing CD4+ T cells against structural peptides of ancestral SARS-CoV-2 virus at baseline, and 7 and 28 days after a randomised third dose of CoronaVac or BNT162b2 vaccine in adults who previously received two doses of either vaccine.** Data was available from a random subset of 20 participants selected from each study arm. The four study arms included participants who previously received two-dose CoronaVac and were randomised to receive a third dose of CoronaVac ("CC-C") or BNT162b2 ("CC-B"), and participants who previously received two-dose BNT162b2 and were randomised to receive a third dose of CoronaVac ("BB-C") or BNT162b2 ("BB-B"). The dotted lines represent the lower limits of detection based on DMSO controls and participants with values above these thresholds are considered as responders: 0.000039% for $CD4^+$ $IL4^+$ T cells. The numbers above each panel indicate the mean level, while the percentages indicate the proportion of responder. p-values ≤ 0.05 *, ≤ 0.01 **, ≤ 0.001 ***

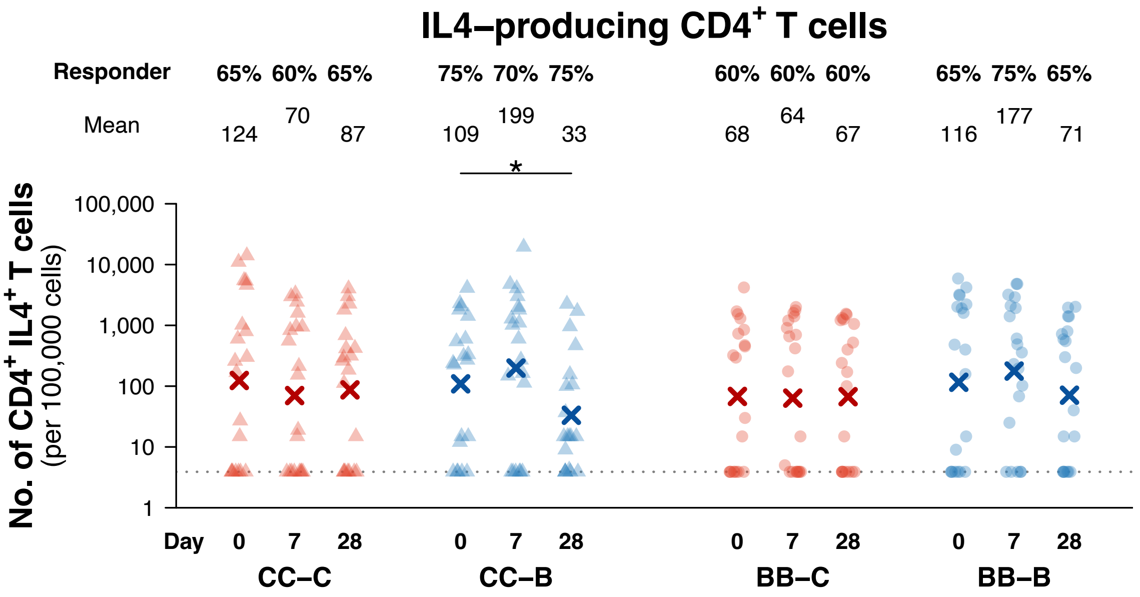
